# Genetic architecture of cardiac structure and function

**DOI:** 10.1101/2024.10.08.24315091

**Authors:** Chang Lu, Kathryn A McGurk, Sean L Zheng, Antonio de Marvao, Paolo Inglese, Wenjia Bai, James S Ware, Declan P O’Regan

## Abstract

The heart is the first organ to develop in the mammalian embryo and its architecture is dependent on the spatial organisation of diverse cell types and morphogenetic transformations shaped by biophysical forces. Cardiac remodelling occurs in the mature heart and is a cascade of adaptations in response to stress that are primed in early life. A key question remains as to the processes that regulate the geometry and motion of the heart, and how it adapts to stress. Here we performed a spatially-resolved genome-wide association study of three-dimensional cardiac traits in 47,549 participants of UK Biobank. We found 42 loci associated with cardiac structure and contractility many of which reveal patterns of spatial organisation in the heart. Newly discovered loci relate to pathways implicated in cardiomyocyte differentiation and chamber development as well as those linked to cardiomyopathies. These findings provide a comprehensive description of the pathways that orchestrate heart development and mechanisms that regulate adaptation in health and disease.

## Introduction

Diverse cardiovascular cell types coordinate to form spatially organised structures of the human heart.^1^ Interactions between biophysical mechanics, morphogenesis and cell fate during heart development shape its three dimensional structure.^2^ Cardiac remodelling occurs in the mature heart and is associated with the development and progression of ventricular dysfunction which is manifest by changes in geometry and contractility that are regulated by mechanical, cellular and genetic factors.^3^ Remodelling complicates many cardiovascular disorders and has an effect across diverse cell types and anatomic domains. During cardiac development exposure to diverse stimuli prime gene expression profiles that drive phenotypic and functional adaptations in later life.^4^ Cardiomyopathies in particular are characterised by diverse morphofunctional phenotypes that relate to the additive effect of common and rare variants on the sensitivity to environmental stimuli.^5,6^ While several candidate genes have been implicated in global structural phenotypes through genome-wide association studies (GWASs),^7,8^ the genetic architecture of cardiac geometry and contractile function, as well as associations with cardiomyopathic states, remain poorly characterised.

The structure and function of the heart can be assessed through detailed mapping of wall thickness throughout the left ventricle as well quantifying regional deformation to assess myocardial contractility. Here we use data from over 40, 000 participants in UK Biobank with cardiac magnetic resonance imaging (CMR) and apply deep-learning computer vision techniques to enable spatially-resolved phenotyping of left ventricular geometry and motion (Figure 1).^9,10^ This provides a three-dimensional time-resolved atlas of the heart to perform genetic association studies across the allele frequency spectrum. We report the effect of environmental and genetic factors on the molecular orchestration of cardiac shape, motion and remodelling in adults. This approach shows newly associated loci only discovered through precision cardiac phenotyping, describes the regional pattern of genetic variants associated with cardiomyopathy, and reveals conserved pathways that regulate multi-organ development.

**Figure 1.**
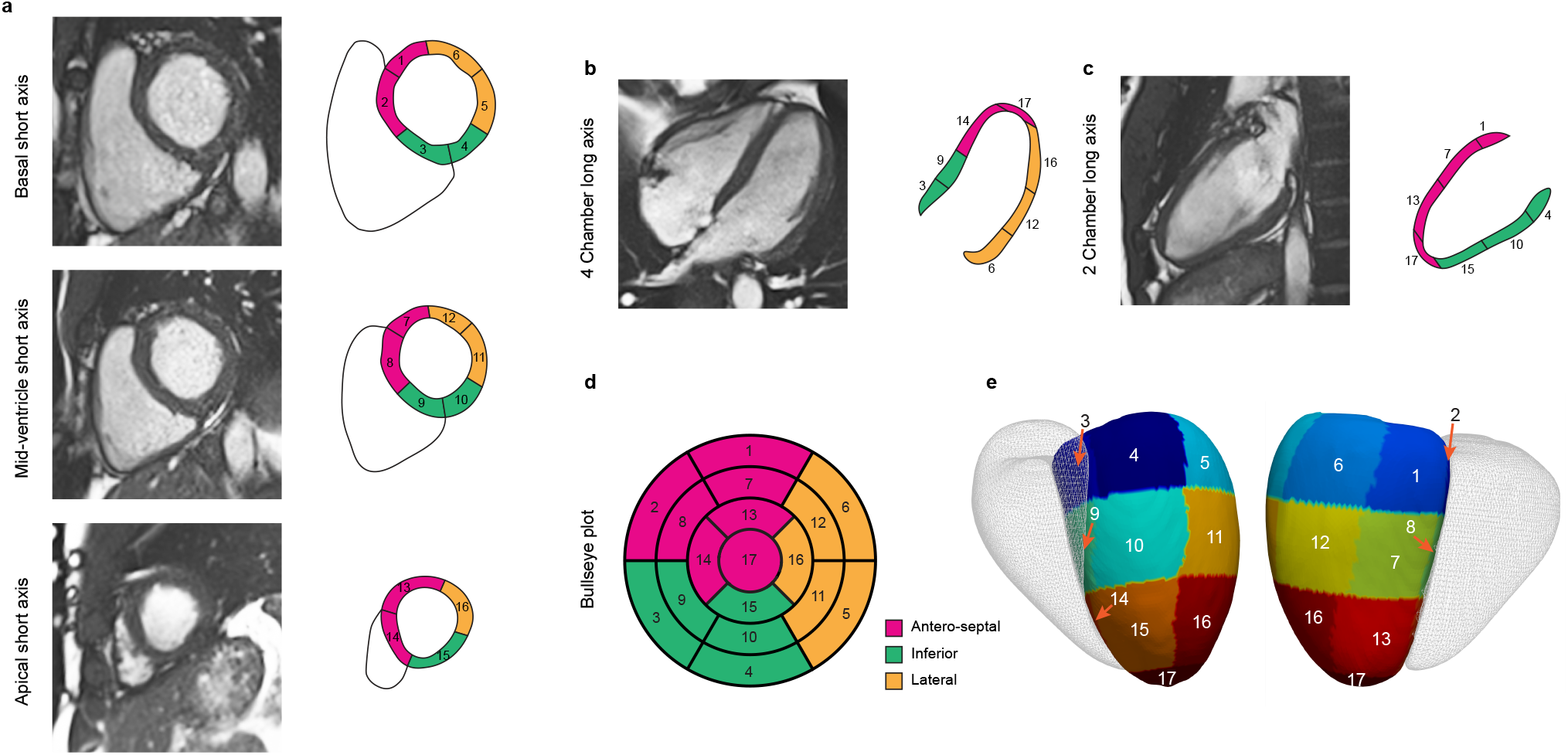
Cardiac image analysis. Cardiac magnetic resonance (CMR) cine imaging was performed in short and long axis planes. A fully convolutional neural network was used to segment the left ventricular myocardium and determine regional wall thickness. Motion tracking was performed using image registration to map myocardial deformation between frames and calculate regional Eulerian strain. The left ventricle was divided into 17 segments using the American Heart Association (AHA) model. Short axis (**a**), four chamber long axis (**b**) and two chamber long axis (**c**) CMR imaging with corresponding segments. Segments are grouped by anatomic location and numbered sequentially allowing characteristics to be represented on bullseye (**d**) and three dimensional (**e**) models of the left ventricle (right ventricle shown in outline).

## Results

### Study overview

We analysed CMR data from up to 47,549 participants of UK Biobank. Deep learning segmentation and motion tracking was used to assess cardiac structure and motion throughout the left ventricle. Data are summarised by anatomical region using the American Heart Association (AHA) 17 segment model (excluding the apex). For each segment mean wall thickness (WT) at end diastole, mean peak circumferential myocardial strain (strain^*circ*^) and mean peak radial myocardial strain (strain^*rad*^) were estimated on short-axis imaging using validated pipelines.^9,11^ In addition, conventional global measures of left ventricular mass and volume were derived.

We assessed patterns of spatial correlation of phenotypes across the left ventricle using biophysical and haemodynamic co-variates (Supplementary Table 1). Phenome wide association studies (PheWAS) were performed on these traits after known confounders had been adjusted for. Genome wide association studies (GWAS), variant-level exome-wide association studies (ExWAS), and gene-level exome-wide burden tests were performed on up to 40,058 healthy individuals of white British ancestry without cardiomyopathy for 54 spatial and global traits. We performed Mendelian randomisation (MR) to analyse potential causal associations between haemodynamic factors and regional wall thickness. To understand the relationship between genetic factors regulating spatial physiology and the patho-physiology of cardiac remodelling we performed genetic correlation and causal association analysis with hypertrophic (HCM) and dilated (DCM) cardiomyopathies.

### Spatial correlations in left ventricular traits

To understand potential drivers of regional phenotypic variation in the heart we assessed spatial correlations in wall thickness and strain across the left ventricle. There was a strong correlation of unadjusted wall thickness between segments, apart from in the basal septum which showed a moderate correlation (Figure 2a to d). Sex, body surface area (BSA), and blood pressure were significantly associated with wall thickness globally (Supplementary Figure 1). After adjustment for these parameters the spatial correlations in wall thickness were reduced to low or moderate (Figure 2g) suggesting other unmeasured factors contribute to variation in hypertrophy.

**Figure 2.**
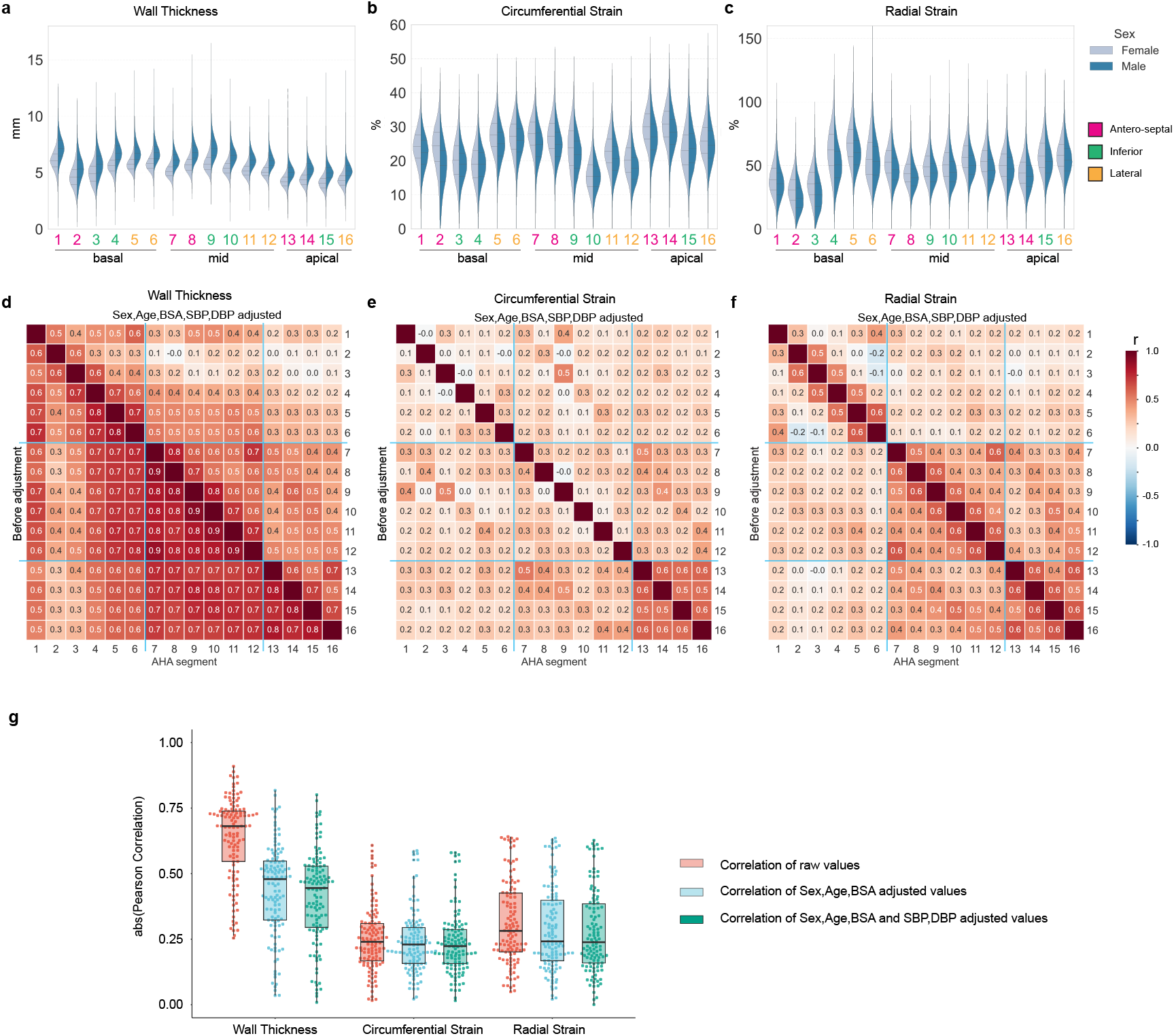
Correlations between spatial left ventricle (LV) traits and association to known predictors. (**a-c**) Distribution of regional traits in up to 47,549 participants by sex are shown with violin plots. Scott’s rule was used to determine the smoothing bandwidth for the kernel estimation. Inner dotted lines show the data quartiles. (**d-f**) Pearson’s correlation among the 16 regional wall thickness (WT), strain^*circ*^, and strain^*rad*^ before (lower diagonal) and after (upper diagonal) adjustment by the indicated confounding factors. The correlation coefficients are shown as heatmaps. The color indicates a correlation efficient from -1 to 1 (blue to red). The basal, mid-cavity and apical segments are separated by light blue lines. (**g**) Distribution of regional trait Pearson’s correlations before and after adjustment by the indicated confounding factors. The boxes show the quartiles of the dataset, and all values are overlaid as dots.

Blood pressure has a significant genetic correlation (*r*_*g*_) with mid-ventricular wall thickness.^8^ Given the high correlation between both systolic (SBP) and diastolic blood pressure (DBP) with wall thickness, we compared *r*_*g*_ of blood pressure with SBP and DBP-adjusted wall thickness (Supplementary Data File 1). We found that *r*_*g*_remained significantly associated with SBP (*P* < 0.003, [*r*_*g*_, 0.13 − 0.27]) for wall thickness across the left ventricle, and with DBP (*P* < 0.04, [*r*_*g*_, 0.09 − 0.20] (Figure 3a and b) for all regions except the basal septum, suggesting that there are shared genetic factors that regulate blood pressure and regional wall thickness independent of a hypertrophic stimulus-response. There is also a significant degree of *r*_*g*_ between HCM and BP-adjusted spatial wall thickness (*P* < 0.024, [*r*_*g*_, 0.19 − 0.56]) (Figure 3c).

**Figure 3.**
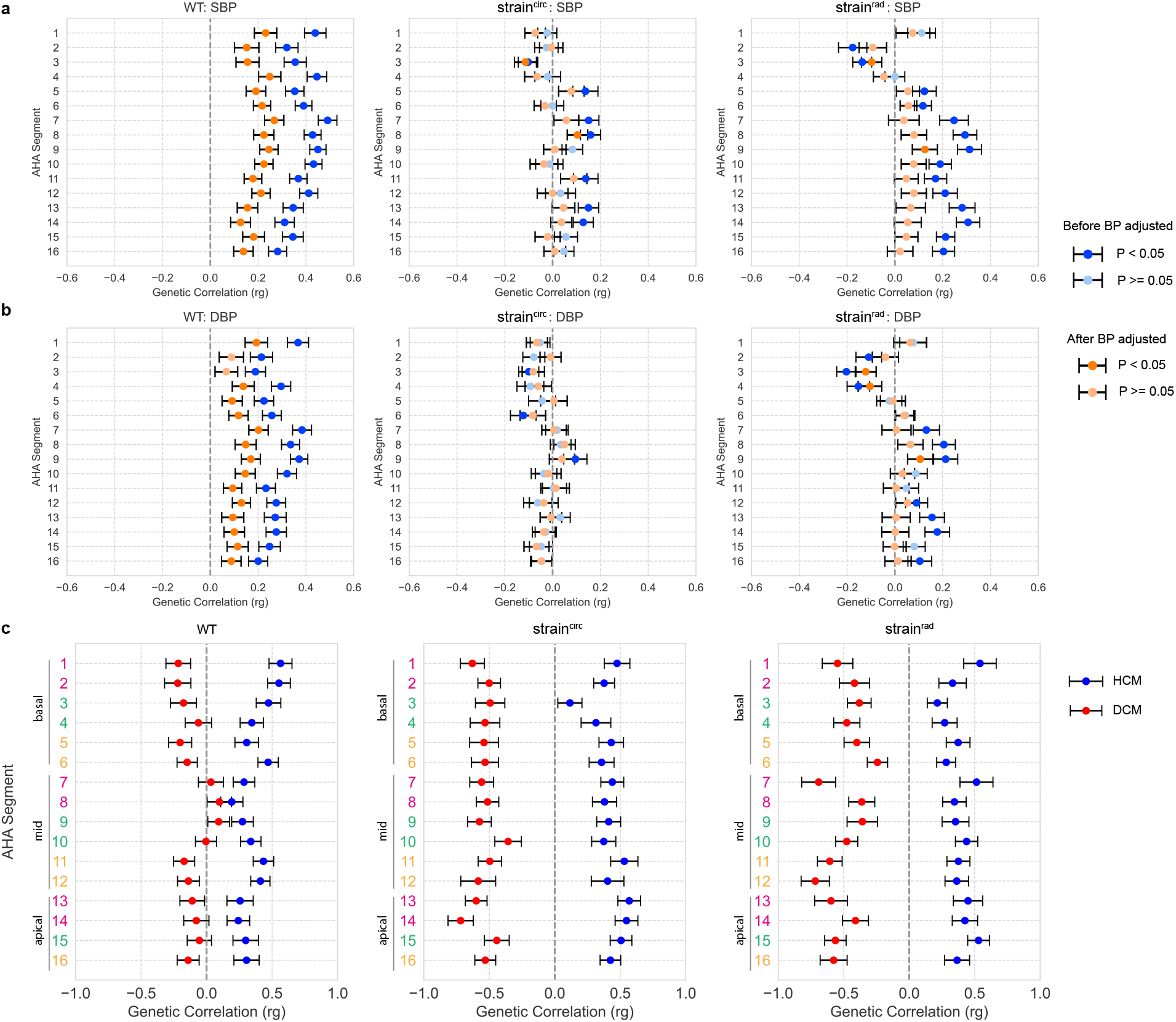
Genetic correlation (*r*_*g*_) between spatial left ventricular traits, blood pressure and cardiomyopathies. The *r*_*g*_ between spatial traits and both systolic blood pressure (SBP) (**a**) and diastolic blood pressure (DBP) (**b**) were calculated before (blue) and after (orange) adjustment for BP at the imaging visit. GWAS of spatial left ventricular traits was performed on up to 40,058 UK Biobank participants of white British ancestry. Summary statistics of SBP and DBP came from a GWAS on up to 801,644 individuals.^14^ Two-sided *P* values were estimated using linkage disequilibrium score regression. Significant correlations (*P* < 0.05) are shown in darker colors. Center values are the estimated *r*_*g*_ and error bars indicate standard error. (**c**) *r*_*g*_ between BP-adjusted spatial traits and hypertrophic cardiomyopathy (HCM) (blue) and dilated cardiomyopathy (DCM) (red). Summary statistics of HCM came from GWAS of 1,733 cases and 6,628 controls,^15^ and DCM from GWAS of 5,521 cases and 397,323 controls.^16^

We observed weaker spatial correlations between segments for myocardial strain (Figure 2e,f), demonstrating more regional independence in systolic function which would not be captured through global parameters. In contrast to wall thickness, known co-variates (sex, age, BSA, BP) have different patterns of correlation with regional strain (Supplementary Figure 1). For instance, BP is associated with decreased strain^*rad*^ in the basal septal wall but increased strain^*rad*^ in mid-ventricular and apical segments (Supplementary Figure 2), and also has opposing directions of genetic correlation (Figure 3a,b). This heterogeneity may reflect anatomic gradients in biomechanical function related to myofibre architecture in the left ventricle.^12^ GREML analysis shows that myocardial strain has lower heritability (*h*^2^, 0.06 − 0.24) than wall thickness (*h*^2^, 0.14 − 0.30), and more unaccounted variance from known co-variates and common SNPs combined (Supplementary Table 2, Supplementary Data File 2). Myocardial strain also has a significant genetic correlation with both HCM and DCM across multiple left ventricular segments (Figure 3c), highlighting shared mechanisms regulating myocyte performance in health and disease.

Phenome-wide association studies of spatial traits show both global and anatomically localised patterns of association with 1,840 phenotypes (Supplementary Figures 3 to 5, Supplementary Data File 3). For instance, hypertension is associated with left ventricular hypertrophy and decreased strain in the basal septum. Heart failure shows widespread impairment of strain while coronary atherosclerosis was associated with regional changes in both strain^*circ*^ and strain^*rad*^. Spatial associations in structure and function were also related to conduction disorders and arrhythmias, as well as cardiometabolic diseases. Genetic variants are known to be associated with cancer therapy–induced cardiomyopathy and we found that chemotherapy was associated with regionally reduced strain without hypertrophy.^13^

### GWAS of spatial cardiac traits

We investigated which common genetic factors underlie regional structural and functional traits in the left ventricle. We used a conservative significance level (*P* < 3.125 × 10^−9^) based on the conventional genome-wide significance threshold (5 × 10^−8^) to adjust for the multiple tests that were performed for each trait spatially. At this threshold, 42 genomic loci were discovered for the three image-derived spatial traits comprising 21 loci for wall thickness, 16 for strain^*circ*^ and 18 for strain^*rad*^ (Figure 4a and Supplementary Table 3). Gene prioritisation (Supplementary Data File 4) was performed by considering the nearest gene, and tissue specific annotations by expression quantitative trait loci (eQTL) and chromatin interactions (CI) in the left ventricle, aorta, and other artery-related tissues using FUMA.^17^ The majority (>90%) of genes or gene products were annotated by location and the left ventricle specific eQTL (Supplementary Figure 6). Mendelian cardiac genes in the GWAS loci were mapped using the Cardiac Genotype-to-Phenotype (CardiacG2P) database (Supplementary Table 4).^18^

**Figure 4.**
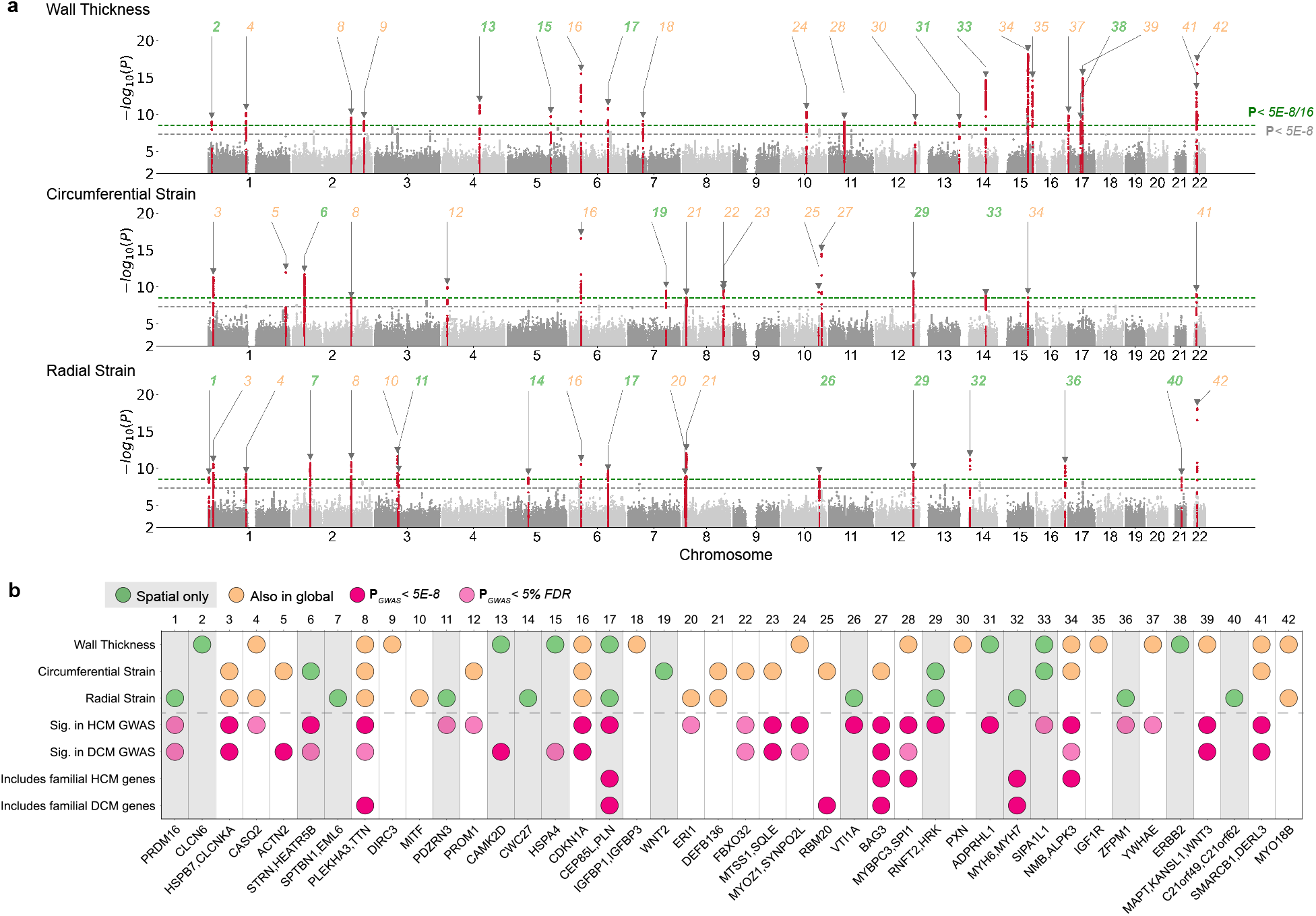
GWAS of spatially resolved wall thickness and contractility. (**a**) Manhattan plots for spatially resolved analysis on three left ventricular traits, namely wall thickness, strain^*circ*^ and strain^*rad*^. The plots show variant-based two-sided minimum *P* values from 16 GWAS of left ventricular AHA segments for the indicated traits for up to 40,186 Europeans in UK Biobank. In the merged spatial analysis, each locus was defined at above multiple-hypothesis adjusted genome-wide significance threshold of 3.125 × 10^−9^ (green dotted line) and labelled sequentially by chromosome location across all loci. The conventional genome-wide significance at 5 × 10^−8^ is shown as a grey dotted line. Spatial-only loci are labelled in bold green and those that were found significant in global traits by conventional genome-wide significance were labelled orange. Locus 8, 28, 29, and 39 contain more than one independent lead SNP (LD r^2^ < 0.1). (**b**) Locus look up in cardiomyopathy genes. The loci not significant in the corresponding globally averaged trait are highlighted with a grey background. Significance is defined as having at least one SNP in the corresponding locus window that reached conventional genome-wide significance (dark pink) or 5% FDR (light pink) for the indicated cardiac disease. Locus naming was performed primarily by gene prioritisation considering FUMA and prior gene association with Mendelian HCM or DCM. See Supplementary Table 3 for list of locus genes. Supplementary Figures 7 to 9 show beta values of lead SNPs in the spatial GWAS loci by individual segments.

Eighteen out of the 42 loci identified with spatial phenotyping would not have been discovered through GWAS of conventional global traits of mass and volume (including global mean/maximum wall thickness, global mean strain^*circ*^ and strain^*rad*^, left ventricular mass and ejection fraction) (Figure 4a). Among the 18 loci we found 11 that overlapped with either a hypertrophic or dilated cardiomyopathy locus (Figure 4b).^15,16^ Specifically, these include 6 loci (*STRN, CAMK2D, PLN, VTI1A, TBX3, ADPRHL1*) that reached a threshold (*P* < 5 × 10^−8^) in HCM or DCM GWAS, and an additional 5 loci (*PRDM16, PDZRN3, HSPA4, SIPA1L1, ZFPM1*) that reached a 5% FDR cutoff. One additional locus was mapped to the sarcomeric gene *MYH6*, which encodes the alpha heavy chain subunit of cardiac myosin and is linked to an electrocardiogram GWAS^19^ and congenital heart disease (ClinGen, CCID:008375). Two genes in these loci were linked to Mendelian cardiomyopathies, including *PLN* and *MYH7*, where *MYH7* was mapped through artery aorta eQTL (Supplementary Table 4). The remaining 6 spatial-only loci, that have not yet been found to be directly associated with HCM or DCM, contained genes or gene products related to myocardial wall stress and vascular smooth muscle contractility (*CLCN6*^20,21^), cardiomyocyte differentiation and cardiac development (*SPTBN1*^22^, *WNT2*^23^) and heart failure (*STARD3*^24^). The spatial loci lead SNPs show non-uniform patterns of spatial association to left ventricular wall thickness and contractility (Supplementary Figures 7 to 9). Together, these show that a spatially-resolved GWAS provides additional power to identify genomic variants that are disease relevant.

### Transcriptome-wide association study (TWAS) for gene prioritisation

Cis-eQTL variants significantly associated with protein expression in left ventricular tissue (GTEx v8)^25^ were mapped in 17 of the 42 GWAS loci. We therefore performed eQTL transcriptome wide association analysis (TWAS) to assess the potential downstream regulatory effects of GWAS variants on expression. TWAS was performed on regional GWAS summary statistics using the MetaXcan framework^26^ and the GTEx v.8 eQTL MASHR-M models (http://predictdb.org/).^25^ We used Bonferroni correction for the number of genes tested to identify significant gene associations. TWAS results are reported for regional wall thickness (Figure 5), strain^*circ*^ and strain^*rad*^ (Supplementary Figure 10).

**Figure 5.**
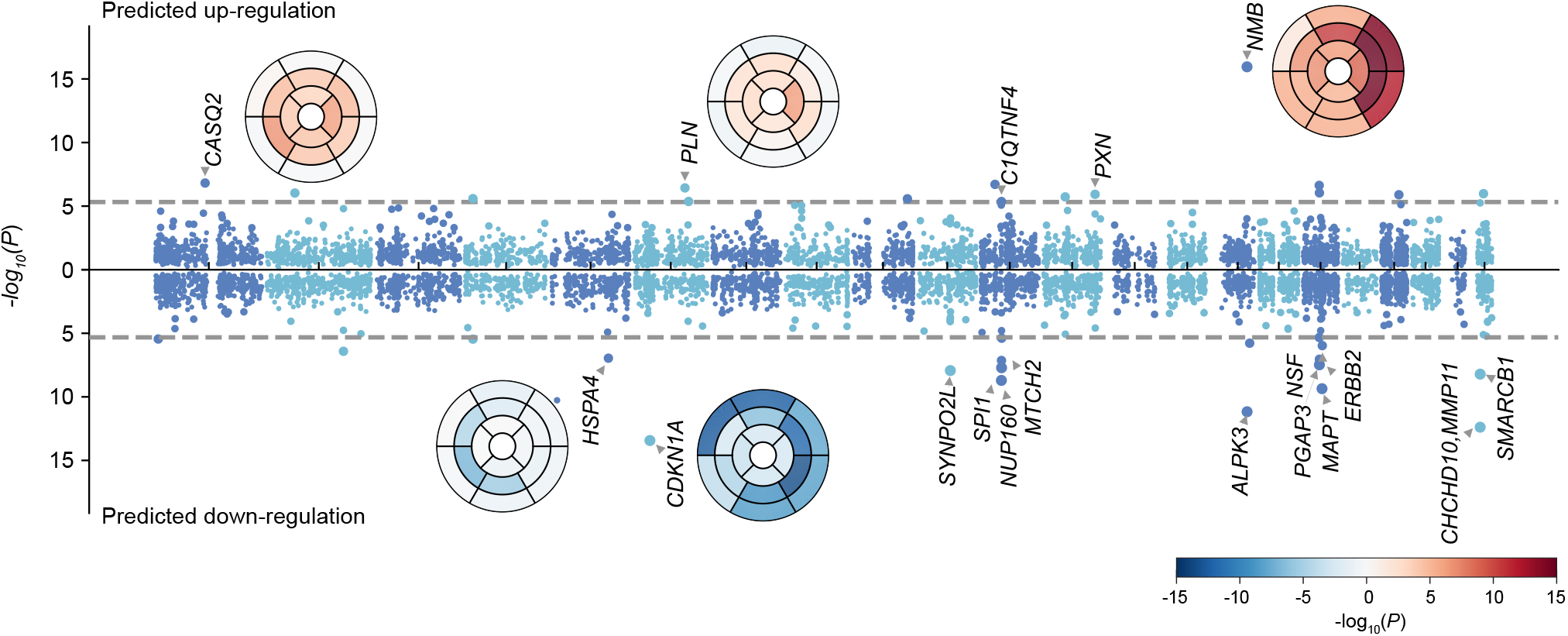
Predicted regulatory effects of GWAS variants on expression. Regulatory effects were calculated using GWAS summary statistics and GTEx v8 eQTL MASH-R model for the “heart left ventricle”. 10,498 genes were tested. Manhattan plot shows the minimum *P* value for segmental wall thickness, with chromosomes coloured by alternate dark and light blue. Bullseye plots were colored with red (positive effect of predicted gene expression on wall thickness) and blue (negative effect of predicted gene expression on wall thickness). eTWAS plots for strain^*circ*^, strain^*rad*^ are shown in Supplementary Figure 10.

We found common variants associated with increased wall thickness and strain rates correspond to significant down-regulation of expression for *CDKN1A, SYNPO2L, ALPK3, MAPT, MMP11* and *SMARCB1*, and to significant up-regulation for *CLCNKA, CASQ2, HSPB7, NMB*. We also observed regional patterns of significance for known global loci with examples (*CASQ2, CDKN1A, NMB, ALPK3*) shown as bullseye plots (Figure 5).

### Variant level exome-wide association study (ExWAS)

Using the whole exome sequencing (WES) data, we tested variant-level association for those not included in the imputed genomes, and for rare variants with ≥7.8×10^−5^ minor allele frequency (MAF) in the CMR cohort. After quality control, 1,006,431 exome variants were included, 73.8% had a MAF < 0.001 and about 25% had an Ensembl impact level of high or moderate (Supplementary Figure 11). At the conservative threshold of *P* < 3.125×10^−9^, 655 significant genotype-to-spatial trait associations from 154 exome variants were identified, of which 82 associations were from 19 exome variants not identified at GWAS (listed in Supplementary Table 5). Overall, ExWAS identified variants in 20 of the 42 GWAS loci, and 3 additional variants outside the GWAS loci (Figure 6). An example of these additional variants was a rare missense variant in *CSRP3* (rs45550635, MAF 0.5%, missense W4R), which associated with increased strain^*circ*^ and left ventricular ejection fraction. *CSRP3* is a cardiomyopathy-associated gene implicated primarily with HCM, but has not been discovered in previous GWAS. These ExWAS summary statistics were combined with GWAS results to provide a comprehensive interpretation of associated variants.

**Figure 6.**
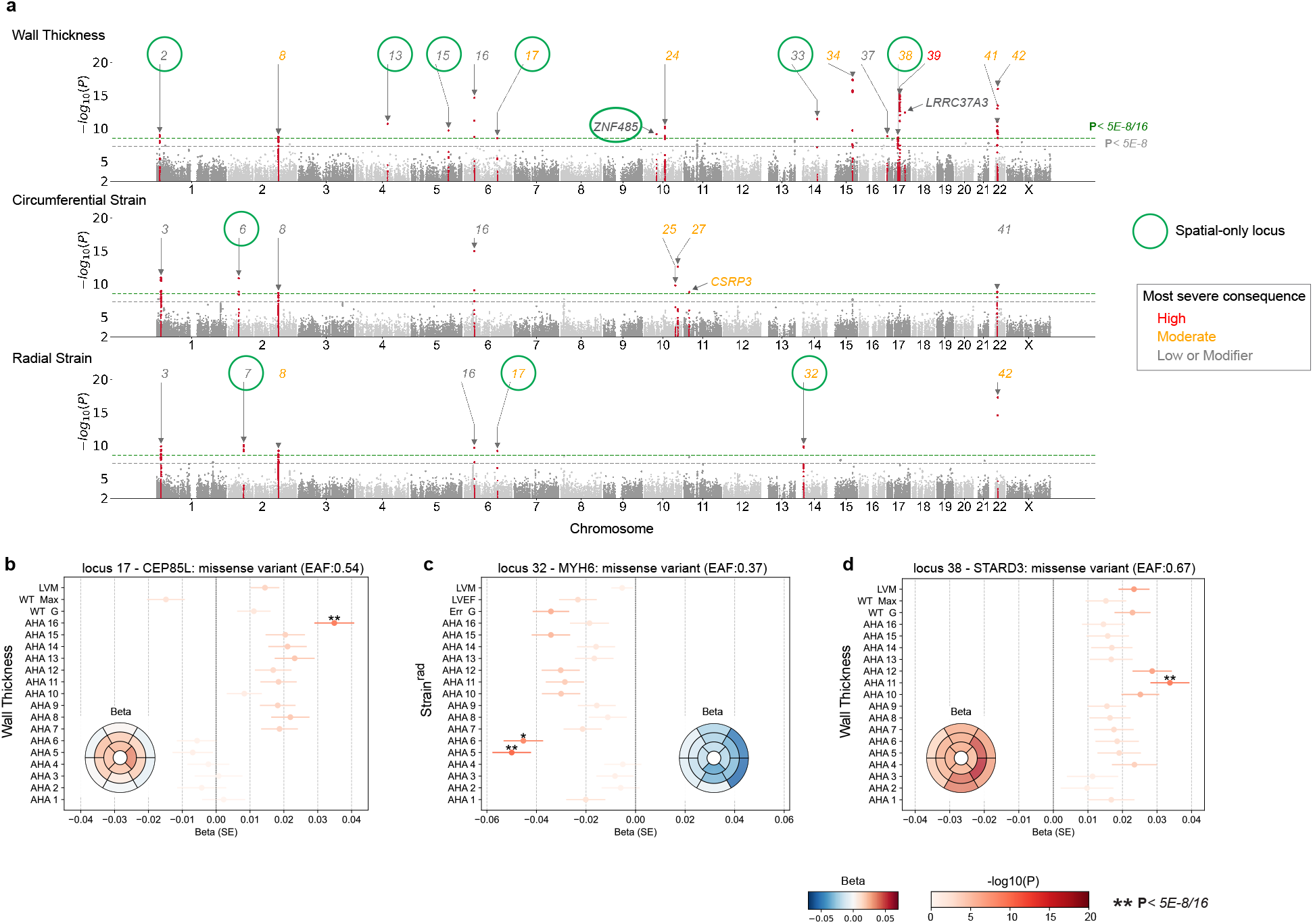
Exome-wide variant-level association study (ExWAS) on spatially resolved wall thickness and contractility. (**a**) Manhattan plot of ExWAS for spatial traits. The smallest *P* values across 16 segments were plotted for wall thickness, strain^*circ*^ and strain^*rad*^. Locus definitions were taken from the GWAS analysis. Three exome variants that reached significance but not mapped to the 42 GWAS loci are labeled with the gene names. The loci are coloured by the most severe consequence of the variants. Spatial only loci from the GWAS analysis are circled in green. (**b**) Forest plots to show the association beta and *P* value for selected variants on spatial wall thickness, global mean wall thickness (WT G), maximum wall thickness (WT Max) and left ventricular mass (LVM). Forest plots are coloured by log-scaled *P* values and significant ones are labeled with asterisks. The insets are bullseye plots of beta values. Direction of beta values are specific to the effect alleles. The variants plotted are (in GRCh38): chr6:118566140:T:C (CEP85L), chr14:23392602:A:G (MYH6), chr17:39657827:G:A (STARD3), from left to right.

We analysed ExWAS variants within the 42 loci identified in the spatial left ventricular trait GWAS. There were 6,631 exome variants tested in the 42 loci, of which 5,683 were not included in the imputation set GWAS was performed on. We found that 87.1% of these exome-only variants were rare (MAF < 0.1%), and 96.3% of those with high impact were also rare (truncating, splice donor or splice acceptor) (Supplementary Figure 11f). These mostly rare and high impact exome variants were observed in 14 GWAS loci, of which 10 were also HCM or DCM loci that include stop gained variants in genes (*CLCNKA, TTN, CEP85L, PRAG1, AGAP5, SYNPO2L, ADPRHL1, ALPK3, LRRC37A2, SMARCB1*), listed in full in Supplementary Table 6. The majority of these variants were very rare (at the limit of observed at least five times in the cohort, MAF < 0.01%), and had a larger effect size than GWAS significant lead variants. This catalogues functionally significant variants in reported cardiomyopathy-related genes observed in a population that excluded those with diagnosed cardiomyopathy.

A spatial approach to phenotyping identified loci containing Mendelian genes implicated in cardiac diseases that did not reach significance using global traits (Figure 6). In the *CEP85L*/*PLN* locus, the sentinel variant (rs3734381) has been identified from previous HCM and QRS duration GWAS^27^ but has never been identified for left ventricular wall thickness or mass. Here, we found this variant to be associated with spatially resolved wall thickness with an apical to basal gradient (Figure 6b). In addition, a missense variant in *MYH6* (rs365990) was not found associated with left ventricular mass, ejection fraction or global strain^*rad*^, however, it has a spatial strain^*rad*^ association in the basal lateral wall (β = -0.05, *P* < 3.125×10^−9^, Figure 6c).

### Exome-wide gene-based tests on rare deleterious variants

We further performed gene-level burden tests to assess the collective effect of rare loss-of-function or deleterious variants which were too rare to be tested at variant level. Using masks provided by UK Biobank^28^, up to 9,865 genes were tested for predicted loss-of-function (pLoF) variants, and 15,615 genes for deleterious variants using Regenie^29^ (see Methods for details). Variance component tests that takes into account the directionality of effects were also performed on up to 3,666 genes for pLoF variants, and up to 9,732 for deleterious variants. For spatial traits, *P* values of genes tested on all 16 segments by the same test were merged to assess false discovery, and a significance level threshold was set at 5% FDR (q value < 0.05). Supplementary Data File 5 shows full list of genes that passed this threshold.

For genes harbouring rare (MAF < 0.01) pLoF variants, *EXD2* and *MYOT* (*P* < 1.4 ×10^−5^) significantly associated with left ventricular mass, *EXD2* and *PCDHA12* (*P* < 7.8×10^−6^) with global mean wall thickness, and *TTN* with ejection fraction, radial and circumferential strains. Carriers of *EXD2* rare pLoF variants had decreased left ventricular mass and wall thickness (β =-1.1, -0.9 respectively). *TTN* is a definitive-evidence dilated cardiomyopathy gene where truncating variants are associated with cardiac phenotypes.^30^ We also identified *ALPK3* as having a spatial wall thickness association through variance component tests (SKAT, *P* = 9.28×10^−8^) that takes into account different directions of effect from pLoF variants (Supplementary Figure 12a). The LOVO tests showed the direction of effect for each pLoF variant with respect to changes in wall thickness (Supplementary Figure 13).

For genes harbouring rare predicted deleterious variants, we found an association between *INHBB* with spatial wall thickness, and *CSRP3* with spatial and global contractility (Supplementary Figure 12b). *CSRP3* and *TTN* displayed opposite directions of association with contractility: the deleterious variants of CSRP3 were associated with increased strain^*circ*^ while *TTN* pLoF variants were associated with decreased strain^*circ*^. *INHBB* deleterious variants (0.014% of cohort) showed a protective role against increased wall thickness (β = −1.1, *P* = 1.84×10^−6^). We also found an association of *AQP1* with spatial wall thickness by the variance component tests (SKAT, *P* = 3.90×10^−7^). *AQP1*, aquaporin 1, is a water channel protein differentially expressed in cardiovascular tissues where it regulates transmembrane homeostasis and is also associated with cardiovascular outcomes.^31,32^

### Causal association with haemodynamics

Both SBP and DBP have an established causal influence on HCM,^6,15^ and also mediate elevated left ventricular mass in the general population,^33^ although the patterns of remodelling may be distinct in these groups.^5^ Here, we analysed the causal relationship of SBP and DBP on global and spatial traits.

We used MR Egger to test for causality as this method accounts for pleiotropic effects. Both SBP (Egger, *P* = 2.82×10^−12^) and DBP (Egger, *P* = 1.65×10^−8^) have a causal relationship with mean wall thickness, where DBP has a higher effect ratio (Supplementary Figure 14). We further performed the MR analysis of BP on spatial traits and found significant associations at adjusted *P* values (Egger, *P* < 0.003125), and performed reverse MR using spatial traits as exposures and BP as the outcome where no significant reverse causality was found. We used the inverse variance weighted test (IVW) to estimate the magnitude of causal association, and observed a gradient of stronger odds ratio of wall thickness increase per standard deviation change in BP from base to apex, for both SBP and DBP (Figure 7a). This shows a causal relationship between BP and left ventricular geometry.

**Figure 7.**
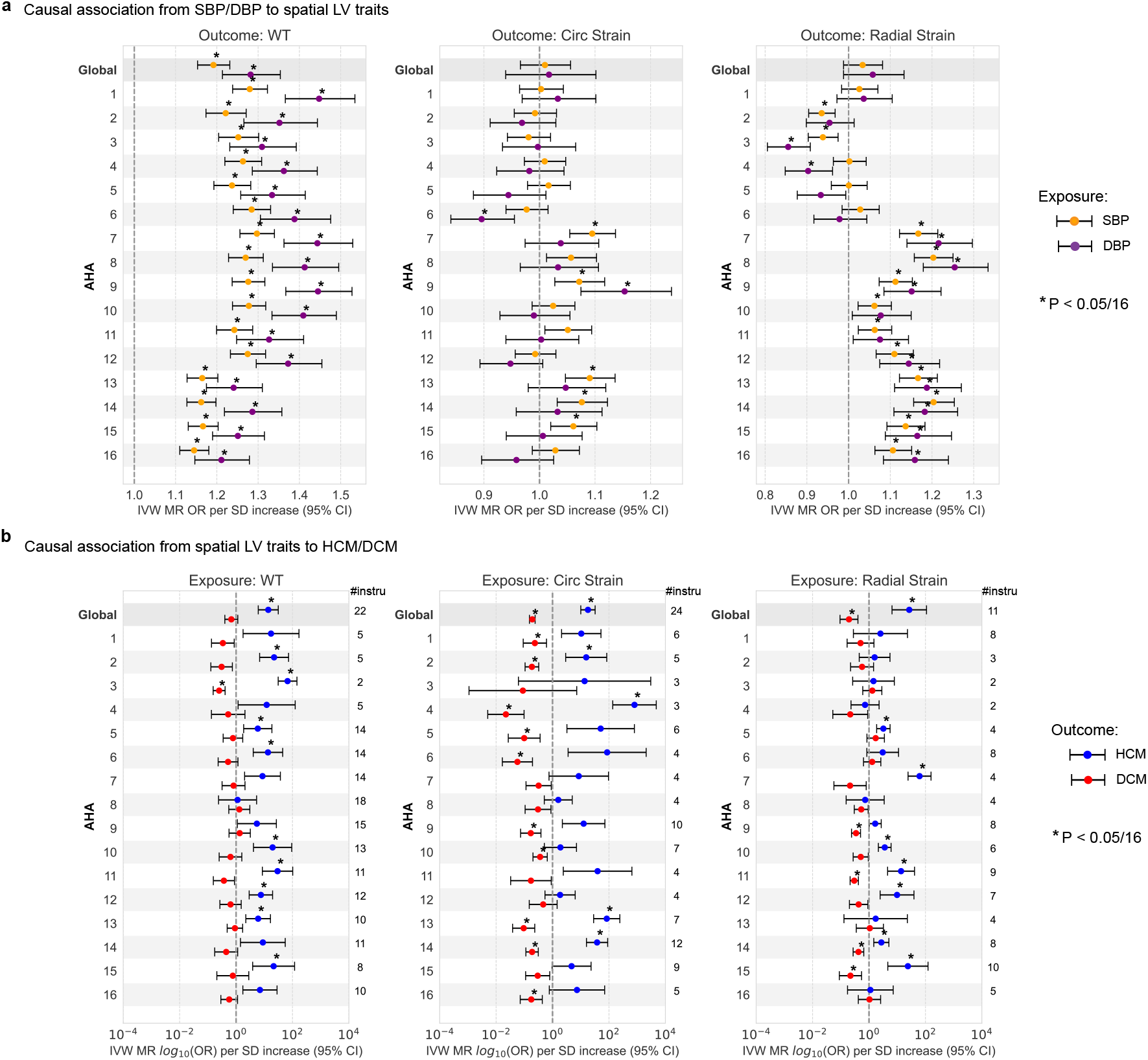
Mendelian randomization (MR) analysis of blood pressure on spatial traits, and spatial traits on the risk of HCM and DCM. Odds ratio (OR) represented are those inferred from the inverse variance weighted (IVW) two-sample MR per standard deviation increase (SD). The error bars represent the 95% confidence interval of the odds ratio (OR). OR for circumferential strain reflect those of increased contractility. Asterisk indicates the MR IVW test *P* value is below multiple hypothesis adjusted threshold (*P* < 0.05/16). (**a**) MR results on increased SBP/DBP on risk of increased spatial spatial traits, including the global mean (G) and regional mean on AHA segments (1-16) for wall thickness, strain^*circ*^ and strain^*rad*^. Spatial traits used GWAS on rank-based inverse transformed (rIVT) adjusted with Sex, age, BMI and BSA at MRI, in 40,058 participants of the UKB without cardiomyopathy and with available CMR. Genetic instruments for SBP, DBP were selected from a published GWAS including up to 801,644 individuals.^14^ (**b**) MR results on increased global and spatial left ventricular wall thickness, strain in circumferential and radial directions on risk of HCM and DCM. Genetic instruments for spatial traits were selected from the present GWAS and the number of SNPs involved in performing MR to HCM are listed on the right side of each panel. The outcome HCM GWAS included 5,927 cases vs. 68,359 controls^15^, DCM included 14,255 cases vs. 1,199,156 controls.^16^

A causal effect of SBP and DBP on myocardial strain was observed regionally even though there was no causal effect on global mean strain^*circ*^ or strain^*rad*^ (Figure 7b,c). Higher SBP and DBP was causally linked to increased strain^*rad*^ in mid antero-septal segments (AHA 7,8, IVW, *P* = 5×10^−5^, 1×10^−6^ for SBP, *P* = 1×10^−4^, 8×10^−5^ for DBP), while the direction of effect was opposite for inferior basal segments. For spatial strain^*circ*^, increased SBP and DBP are related to decreased strain in basal-lateral segments and increased strain in mid-inferior septal segments, although the causal effect is insignificant.

### Causal association of spatial traits and cardiomyopathy

Prior data from GWAS and MR support a causal role of increased global left ventricular contractility in both obstructive and non-obstructive forms of HCM.^15^ We performed bidirectional Two-Sample MR tests between spatial LV traits and HCM, DCM risks (Supplementary Data File 6). Egger’s test showed that contractility was causally associated with DCM, but not with HCM. We found that the IVW test supported a causal role of global and regional mean wall thickness for HCM, as well as global and regional myocardial strain for HCM and DCM risk (Figure 15b). Egger’s test did not provide additional evidence for a causal role of mean wall thickness on HCM or DCM risk, and the high intercept suggested pleiotropic effects (Supplementary Figure 15).

### Pathways associated with remodelling

We next identified potential interactions among protein-coding genes associated with regional traits. We performed unsupervised gene clustering and pathway enrichment analysis on the interaction network using the STRING-DB server (Supplementary Data File 7).^34^ Protein-coding genes that were significant in global mean wall thickness were clustered into three groups (Figure 8a). The largest cluster contained 10 genes (*WNT3, NDUFS3, MTCH2, MAPT, NSF, CRHR1, KANSL1, PLEKHM1, ARHGAP27, CELF1*) that were significantly enriched for brain volume and cortical area, and 3 genes (*PXN, IGF1R, YWHAE*) with enrichment for left ventricular mass to end-diastolic volume ratio. These genes together also constitute a significant enrichment of anthropometric measurements (Figure 8c). The other two clusters have enrichment in heart and muscle development (*ALPK3, SYNPO2L, MYO18B, MYBPC3, TTN*), and reported GWAS on image-derived traits (*NMB, WDR73, ZNF592*).^7^ The enrichment analysis of mean wall thickness GWAS-identified genes showed processes that are known to affect global development of the left ventricular wall. The re-discovery of genes underlying brain and anthropometric measurement shows genetic components that underlie muscle and size development have shared influence across human organs.

**Figure 8.**
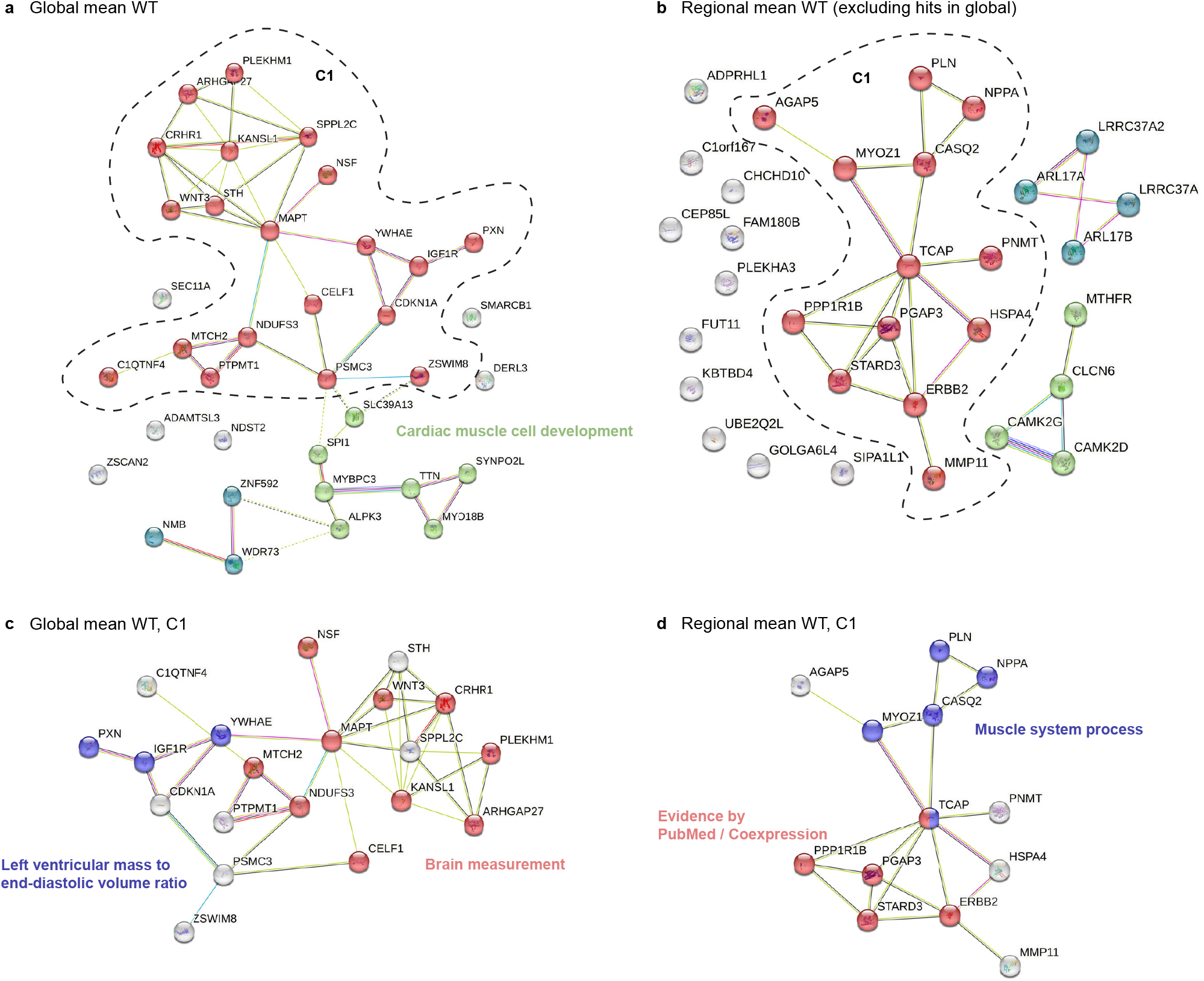
Cluster groups and gene interactions for left ventricular wall thickness. (**a**) Genes in the global mean wall thickness GWAS loci were clustered by the STRING-DB. Enrichment analysis on the largest cluster is shown in (**c**). Edges show protein–protein relationships where a protein works together to perform a common task. (**b**) Genes in the spatial-only loci of regional wall thickness were clustered by the STRING-DB. Enrichment analysis on the largest cluster was shown in (**d**). Light blue line: known interactions from curated datasets; purple line: experimentally determined known interactions; dark blue line: predicted interactions from gene neighborhood; red line: predicted interactions from gene fusions; dark blue line: predicted interactions from gene co-occurrences; light green line: predicted interaction from text mining; black line: co-expression; light purple: gene homology.

Protein-coding genes that were significant in spatial-only loci for wall thickness were clustered into three main groups with 11 genes un-clustered, suggesting the pathway information behind the spatial-only hits is less understood (Figure 8b). The largest cluster identified was enriched for muscle system process genes and a cluster centered on *STARD3* (Figure 8d). The StAR Related Lipid Transfer Domain Containing 3 (*STARD3*) gene was reported to contain deleterious or damaging protein-coding variants for heart failure with reduced ejection fraction.^24^

We further included genes in spatial-only loci for strain^*circ*^ and strain^*rad*^ in the analysis. These include 84 protein-coding genes identified from the 18 spatial-only loci, and STRING analysis identified 7 clusters (Supplementary Figure 16). Twenty two genes constitute the largest connected cluster, involved in the sarcomere and heart development (*MYH6, MYH7, PLN, ERBB2, WNT2, TCAP*), and the ERBB2 pathway (*STAT5A, STAT5B, HSPA4, NPPA*). The other clusters identified genes involved in the regulation of excitation contraction coupling in heart and voltage-gated chloride channels (*CAMK2D, IFNGR2, UQCRQ*), genes associated with splicing regulation (*SREK1IP1, CWC27, PPWD1*), genetics associated with cardiac QRS duration and cardiac ventricular conduction (*HEATR5B, STRN*).^27^ This analysis provides an extended view of genes and pathways involved in the regulation of cardiac physiology.

## Discussion

The three dimensional structure of the heart in health and disease shows high endophenotypic diversity which is predictive of outcomes.^5,35,36^ Here, we aimed to discover the genetic determinants of patterns of structural and functional adaptation in the heart through spatially-resolved phenotyping and genetic association analyses. Using deep learning derived anatomic atlases we found spatially-distinct associations in loci related to cardiac chamber development and stress-response pathways. We also discover novel associated loci not detected using global traits, identify potential pathways that regulate phenotypic adaptation, and dissect the causal relationships between environmental stimuli and remodelling. Together, this work provides a comprehensive resource for understanding the molecular underpinnings of cardiac anatomy and physiology as well as the response to stress.

A spatially-resolved GWAS of left ventricular traits enables the discovery of genetic associations that have regional expression. Newly discovered loci include several that are associated with cardiomyopathies in case-control GWAS for HCM (*CEP85L*/*PLN, ADPRHL1, SIPA1L1*) and DCM (*HSPA4, CAMK2D*), as well as the sarcomeric gene *MYH6*, which is linked to congenital heart disease but has not previously been associated in left ventricular global trait GWAS. Spatial contractile function reveals 2 loci (*STRN, PRDM16*) that are also found in cardiomyopathy GWAS. *STARD3* is not related to cardiomyopathy but is associated with heart failure. *PLN* and *MYH7* (eQTL hit in the *MYH6* locus) were both reported as pathogenic genes in hypertrophic cardiomyopathy.^37^ These findings extend the known repertoire of shared genetic loci between cardiac development and adaptation in adults with GWAS and Mendelian cardiomyopathy-associated genes. Exome-wide analyses discovered a further 3 loci including a missense variant in *CSRP3* (amino acid mutation W4R) which was analysed in cardiomyopathy Mendelian studies but not yet reported in GWAS catalog, highlighting the additional power for variant-level discoveries against using impute-only data. Gene-level burden tests looked at the effect of ultra-rare pLoF variants. Rare loss-of-function variants in the cardiomyopathy gene *ALPK3*, which regulates cardiomyocyte and myofibroblast differentiation,^38,39^ were associated with increased regional wall thickness. *ALPK3* is one of several reported genes that have a recessive association with cardiomyopathy supporting a semi-dominant model of inheritance.^40^ We found that common missense variants in *ALPK3* are associated with decreased wall thickness, whereas the pLoF variants are together associated with hypertrophy suggesting a potentially protective role for GWAS variants against hypertrophy. Similarly, the sentinel missense variant in *MYH6* was found to be associated with decreased basal wall thickness, whereas predicted damaging variants are associated with increased wall thickness in cardiomyopathy.^41^

Blood pressure has a key causal influence on global left ventricular hypertrophy with early involvement of the basal septum.^9,42^ Non-sarcomeric HCM is also characterised as a polygenic hypertrophic sensitivity to DBP.^6^ We showed there was a spatially-varying gradient of causal association between blood pressure and wall thickness from base to apex with DBP having a stronger effect. While blood pressure is the causal exposure the hypertrophic response depends on wall stress which is highly heterogeneous in the left ventricle and greatest where the septum is flattest near the outflow tract.^43^ We also found a genetic correlation between physiological variation in hypertrophy and inherited cardiomyopathies -with opposite directions of effect in HCM and DCM. Causal analyses show how common genetic variation modifies Mendelian disease risk and morphofunctional expression contributing to the diversity of cardiomyopathy phenotypes.

Using unsupervised gene clustering and pathway enrichment analysis we found 10 genes that share an enrichment for brain volume and cortical area. There is a shared genetic influence between markers of heart and brain health,^44^ and our findings suggest some genes may contribute to multiorgan development and the emergence of complex traits. Genes discovered only through spatial phenotyping revealed three main groups with the largest regulating heart regeneration and trabecular development through promotion of cardiomyocyte de-differentiation and proliferation.^45,46^ The NRG-1/ErbB signaling pathway is involved in diverse aspects of cardiomyocyte biology has also been identified as a therapeutic target for cardiomyopathy and even heart regeneration.^47^ Pathway analysis also identified genes involved in cardiac conduction suggesting that ion channel genes are critical for regulating the contractility of the adult heart as well as formation of the conduction system.^48^

Together, the data reported here, combining advanced three dimensional human imaging with genetic analyses across the allele frequency spectrum, provide a resource for understanding the mechanisms that govern cardiac development and adaptation as well as contribute to the expression of cardiomyopathic phenotypes.

## Methods

All analyses scripts in this study are available online (https://github.com/ImperialCollegeLondon/spatial-lv-GWAS) and were conducted with Python v.3.11. Genetic analysis were performed on the UK Biobank Research Analysis Platform (RAP).

### Dataset

The UK Biobank^49^ (http://www.ukbiobank.ac.uk) is a large study of about half a million individuals who were recruited across the UK from 2006 to 2010 and for whom genetic and extensive phenotype information has been recorded. All participants provided written informed consent for participation in the study, which was approved by the National Research Ethics Service (11/NW/0382). Our study was conducted under terms of access approval numbers 28807 and 40616. A range of available data were included in this study, comprising genotyping arrays and whole exome sequencing (WES), cardiac imaging, and non-imaging phenotypes. These included sex (p31), age (p21003), height (p50), weight (12143), systolic blood pressure (SBP, p4089) and diastolic blood pressures (DBP, p4080). Specifically, the age, weight, and blood pressure measurements were selected by the instance ID of CMR visit. At the time of analysis, there were 502,396 genotyped participants and up to 47,825 participants who have had cardiovascular magnetic resonance (CMR) imaging. 3168 out of the 440,005 UKB participants who have ICD10 records have been diagnosed with cardiomyopathy, which includes DCM (ICD10 codes: I420, I426, I427), HCM (I421, I422) and anyone else with I42 annotation.

The dataset for genetics association analysis on the Caucasians were selected with the following minimum requirements. Caucasians were selected by UKB genetic ethnic grouping (p22006), and those with sex chromosome aneuploidy (p22019), have outliers for heterozygosity or missing rate (p22027), without imputation data (p22828), or have inconsistent sex (p22001 not consistent with p31) were excluded, reducing the genotyped participants for analysis to 408,098. Overlap of these individuals and those with CMR imaging was a cohort of 40,186, of which 128 have been diagnosed with cardiomyopathy by ICD10 records (demographics in Supplementary Table 1). GWAS analysis were performed on the 40,058 individuals without cardiomyopathy diagnosis. Among the covariates tested, automated blood pressure readings were the largest set of missing data (6800 missing systolic or diastolic readings), therefore when they were considered as covariates, the GWAS were effectively performed on up to 33,158 individuals.

### PheWAS analysis

Phenome-wide association studies (PheWAS) were undertaken using the PheWAS R package with ICD coded clinical outcomes and phenotypes converted to 1,840 categorical PheCodes. *P* values were deemed significant with Bonferroni adjustment for the number of PheCodes.

### CMR imaging protocol and data analysis

A standardised CMR protocol was followed to acquire two-dimensional, retrospectively-gated cine imaging on a 1.5T magnet (Siemens Healthineers, Erlangen, Germany),^50^ and the images all underwent standard quality control prior to use in analysis.^9^ As previously described,^10,51^ automated segmentation of the short-axis and long-axis cine images was performed using fully convolutional networks.^9^ Briefly, left ventricular end-diastolic, end-systolic and stroke volume (LVEDV, LVESV, LVSV) (units in mL) and ejection fraction, (LVEF, %), were determined. Myocardial volumes were used to compute left ventricular myocardial mass (LVM, g) assuming a density of 1.05*g*.*ml*^−1^. Myocardial wall thickness (WT, mm) measurements were derived from automated segmentation. Circumferential and radial strains were calculated using short axis cines as 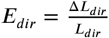, where *dir* represents circumferential or radial direction, *L* _*dir*_ the absolute length of a line segment along this direction and Δ*L*_*dir*_ its change in length over time. The heart was divided into 16 standardised anatomical segments following the AHA protocol, excluding the true apex.^52^ Outlier wall thickness values (>20mm) were manually evaluated by checking the underlying CMR image, and erroneous values were removed.

### Genetic association analysis

Genome-wide association studies, exome-wide association studies, and exome-wide rare variants analyses were all performed using Regenie(v3.1.1),^29,53^ which fits a whole-genome regression model for quantitative and binary phenotypes on a set of high-quality common SNPs (HQ SNPs set), to account for sample relatedness and population structure. All quantitative traits were inverse-rank normalised.

### HQ SNPs set

We selected up to 40,058 healthy individuals for whom CMR data were available and for whom genotyping data passed standard quality checks (UK Biobank Resource 531), namely, excluding those with sex mismatch, close relatives (22021), non-Caucasian (22006), sex chromosome aneuploidy (22019) and outliers for heterozygosity or missing rate (22027) and who have withdrawn participation. We further applied quality-control filters on the genotype data following Regenie methods.^29^ Specifically, the filters were applied using PLINK2^54,55^ (version v2.00aLM, https://www.cog-genomics.org/plink2) and included: a minor allele frequency of > 1%, a Hardy-Weinberg equilibrium (HWE) test not exceeding 1 × 10^−15^, a genotyping rate above 99%, not present in high inter-chromosomal LD, in the major histocompatibility (MHC) region, or in regions of low complexity, not involved in inter-chromosomal LD (LD pruning of 1,000 variant windows, 100 variant sliding windows and *r*^2^ < 0.9) and in chromosomes 1-22. This resulted in up to 519,697 high-quality genotyped SNPs.

### Genome-wide association study (GWAS)

GWAS was performed on 8 million imputed genotypes that passed quality controls, including minor allele frequency (MAF) > 0.5%, imputation score > 0.4, minor allele count > 10, HWE P value > 1 ×10^−15^, and genotyping missing in < 1% individuals (7,955,591 SNPs). Quantitative imaging traits were converted to z-scores using rank-inverse-based normal transformation. Body surface area (BSA) was calculated with the Mosteller formula 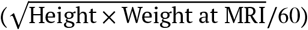. Mean arterial pressure (MAP) was calculated as (2 × SBP + DBP)/3.

In the genome-wide association tests of individual variants, we included covariates: age (age when CMR image was obtained), age squared, sex, age x sex, the top 10 principal components provided by the UK Biobank to appropriately correct for population stratification, BMI and BSA. We performed GWAS with and without adjustment by SBP and DBP. All GWAS were performed using the Regenie (v3.1.1) software. For each trait, Step 1 was run on the HQ SNPs set to obtain the leave-one-chromosome-out (LOVO) predictors. These were then included as covariates in step 2 GWAS on the quality-controlled imputed data. All association analyses used a 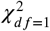 statistic to test a variant for association with a trait (that is *H*_0_ : β_*SNP*_= 0). All programs were called within the UK Biobank research analysis platform (RAP). The results of the regional GWAS analysis on wall thickness, circumferential and radial strain, before and after adjustment by systolic and diastolic blood pressure at MRI are presented in Supplementary Figures 7 to 9.

### GREML analysis

SNP-based heritability were calculated with genome-based restricted maximum likelihood (GREML) implemented in the GCTA software (version 1.93.2beta from biocontainers). GREML were produced using GWAS summary statistics where all traits have variance of 1 after the rank-based inverse variance transform (rIVT). The genetic relationship matrix (GRM) was produced from HQ SNPs set using the GCTA software with grm-cutoff set as 0.039. Fixed effects included the sex, age, BMI, BSA, SBP and DBP measured at MRI. As we performed GWAS using rank-based inverse variance transform (rIVT) of theleft ventricular traits, the total variance is 1 for all traits. From GREML outputs, Vp is the phenotypic variance after fixed effects were accounted for. Thus we defined the percentage of variance accountable by fixed effects as 1-Vp.

### Genetic correlation

We performed pairwise genetic correlation between the 48 spatial LV traits and SBP, DBP, HCM, DCM using LD score correlation (LDSC, v.1.0.1). For each GWAS, summary statistics was prepared using the ‘munge_sumstats.py’ command, filtering for the 1000G European SNPs with corresponding alleles using the –merge-alleles 1000G_eur.snplist flag. We then assessed genetic correlations for each pairs using ‘ldsc.py –rg’, and used the 1000G Phase3 ldscores as reference panel LD Scores. The reference files were downloaded from https://data.broadinstitute.org/alkesgroup/LDSCORE/. We did not constrain the single-trait and cross-trait LD score regression intercepts. The results of the genetic correlation analyses are shown in Figure 3, full results in Supplementary Data File 1).

### Genome-wide significant loci

The genome-wide significant threshold for regional traits (which divided the left ventricle into 16 segments) were selected at *P* < 5 × 10^−8^/16. We defined genome-wide significant loci by iteratively spanning the +/-500 kb region round the most significant variant and merging overlapping regions until no genome-wide significant variants were detected within +/-1Mb, standard specified in^56^ and produced with FUMA.^17^ By this definition, 42 loci were found altogether for the three imaging traits, including 21 for WT, 16 for strain^*circ*^ and 18 for the strain^*rad*^, as shown in Figure 4a, and listed in Supplementary Data File 4. GWASlab^57^ (v3.4.24) was used to make the Manhattan plots. Software for computing q-values from a collection of *P* values is available at: https://github.com/StoreyLab/qvalue.

### Gene prioritisation

Variant annotation was performed using FUMA.^17^ For the 42 spatial GWAS loci, distribution of gene types encoded by Ensembl (v110) by positional variants, those mapped with eQTL variants in “Heart Left Ventricle” or one of the three relevant tissues (Artery Aorta, Artery Tibial, Artery Coronary, and Heart Atrial Appendage) (GTEx v.8^25^), and those mapped by Chromatin Interaction (CI) in “Left Ventricle” or”Aort” tissues (pre-processed significant loops computed by Fit-Hi-C, filtered at FDR 0.05, GEO accession code GSE87112^58^). For eQTL mapping, only significant cis-eQTL pairs (those within 1MB of the gene) were mapped, where significance was defined by the GTEx pre-calculated FDR (gene q-value ≤ 0.05). Distribution of gene types were shown in Supplementary Figure 6. Nominal genes for each locus were limited to protein coding genes (shown in Figure 4), and prioritised by positional overlap, those regulated by eQTL variants in left ventricle, those mapped by CI in the left ventricle and the known Mendelian genes in cardiac conditions from the CardiacG2P database.^18^

### Exome-wide association study

Exome-wide association studies were performed on the final release of whole exome sequencing (WES) data of UKB for the spatial and global left ventricular traits using Regenie (v3.1.1) two-step whole genome regression. The regression model trained on HQ SNPs and the WES data was used to run individual variant associations (minor allele count > 5). After overlap of availability for CMR phenotypes and previous quality checks, the exome-wide associations was run on up to 32,067 individuals (32,067 in spatial WT, 32,053 in spatial strain, 32,065 in LVM, LVEF) and 1,334,918 variants. We further compared the associations results before and after filtering the variants that failed the ‘90 percent have depth above 10’ test (from the ‘500k_OQFE.90pct10dp_qc_variants.txt’ in the UK Biobank helper files), and proceeded only with significant association that passed the test. Finally, the exome variants tested is 1,006,431. Most severe consquences of these variants are annotated using Ensembl Variant Effect Predictor (VEP) functionality with HAIL (HAIL-0.2.78-VEP-1.0.3).

### eQTL and transcriptome-wide association study analysis

We performed transcriptome-wide association using the MetaXcan v0.6.12 tools^26^ and the GTEx v.8^25^ eQTL MASHR-M models (http://predictdb.org/). Each regional GWAS results were harmonized, lifted over to hg38 and linked to the 1000 Genomes Project reference panel using GWAS tools (https://github.com/hakyimlab/summary-gwas-imputation/wiki/GWAS-Harmonization-And-Imputation). Imputed and harmonized GWAS summary statistics were used to perform TWAS for the heart left ventricle in GTEx v.8 with the S-PrediXcan function. Resulting *P* values were corrected using the Bonferroni correction to identify significant gene associations.

### Exome-wide gene-based collapsing analysis

The collapsing analyses on gene-based burden testing was performed on the UKB 470K WES release using Regenie (v3.1.1).^29^ The analyses were conducted on the Research Analysis Platform (https://ukbiobank.dnanexus.com). The same set of regional CMR phenotypes, whole-genome regression model predictors from Regenie step1, and covariates (sex, age at MRI, BSA, SBP/DBP at MRI, and the 10 ethnicity PCs) were used for collapsing analysis. The intersection of the participants in the GWAS analysis, which was performed on the genotyping array, with those having Exome sequencing data is 38,716 (38,583 have regional mean WT observations, 38,563 have the regional mean circumferential and radial strain observations).

Variant annotations (including missense and loss-of-function predictions) were defined by the UKB final release helper file (ukb23158_500k_OQFE.annotations.txt.gz) on the basis of SnpEff annotations from Ensembl gene definitions. We used two masks for variants: (1) predicted loss-of-functions (pLoF), (2) pLoF or missense (5/5), requiring consensus on deleteriousness from 5 scoring algorithms curated in dbNSFP v3.2. Gene based sets were defined by combining the UKB final release helper file (ukb23158_500k_OQFE_c1-22_n0.sets.tsv) and filtered by the 90pct10dp variants list (ukb23158_500k_OQFE.90pct10dp_qc_variants.txt), which for a given variant, requires at least 90% of genotypes, independent of variant allele zygosity, have a read depth of at least 10. In the final masks for pLoF and pLoF+missense, it is required that the minimum minor allele count (MAC) is above 5.

We performed six types of gene-based tests, namely: the additive burden test, the variance component test (SKAT), the test using Cauchy combination method to combine single-variant-*P*-values (ACATV), and the omnibus test combining SKAT and burden (SKATO) and its similar alternative using Cauchy combination to maximimze power (SKATO-ACAT) and the omnibus test combing SKAT, burden and ACATV (ACATO). The number tested for regional WT in the burden mask (ADD) is 10768 for pLoF, 15924 for pLoF+missense, and in the variance component tests is 3666 for pLoF, 9732 for pLoF+missense; similar numbers were tested for the regional strains (only 1-2 less tests performed for the burden masks). Since multiple variance component tests were performed, we reported genes that were significant across all six types of tests in the results.

All raw and derived data in this study are available from UK Biobank (http://www.ukbiobank.ac.uk/), conducted under application number 40616. GWAS summary level data are publicly available through the GWAS catalogue (https://www.ebi.ac.uk/gwas/).For colocalization analyses, we used the unfiltered eQTL results from eQTL Catalogue (https://www.ebi.ac.uk/eqtl/) and the Genotype-Tissue Expression (GTEx) Portal v.8 (https://gtexportal.org/home/).

## Code Availability

The code used for our analyses are available at https://github.com/ImperialCollegeLondon/spatial_cardiac_GWAS.

## Author contributions

C.L. performed the analysis and drafted the manuscript; K.A.McG. performed additional genetic analyses; P.I. performed statistical analysis; W.B. and A. de M. provided the phenotypes; J.S.W. and S.L.Z provided genetic resources for the study; D.P.O’R. conceived the project, provided supervision and funding, and revised the manuscript.

