## Supplementary Information for "Genetic architecture of cardiac structure and function"

#### Supplementary Tables

**Supplementary Table 1. Mean and standard deviation of selected traits.** 47,549 individuals with CMR imaging, and 40,186 Caucasian individuals remained after genotyping quality check. 40,058 individuals with no reported cardiomyopathy were selected for genetic analysis.

| | CMR imaging cohort | Genetic study cohort |
|---|---|---|
| Quantitative trait | Mean (SD) | |
| Age at MRI (year) | 64.3 (7.8) | 64.5 (7.7) |
| Weight (kg) | 75.3 (15.0) | 75.4 (15.0) |
| Height (m) | 169.7 (9.2) | 169.9 (9.1) |
| BMI (kg m-2) | 26.0 (4.3) | 26.0 (4.3) |
| Body surface area (m2) | 18.8 (2.2) | 18.8 (2.2) |
| Systolic blood pressure (mmHg) | 139.5 (18.9) | 139.8 (18.9) |
| Diastolic blood pressure (mmHg) | 78.8 (10.1) | 78.8 (10.1) |
| Mean arterial pressure (mmHg) | 99.0 (11.7) | 99.2 (11.7) |
| LVEDV (mL) | 147.2 (33.9) | 147.7 (33.9) |
| LVESV (mL) | 60.1 (19.5) | 60.4 (19.5) |
| LVSV (mL) | 87.0 (19.3) | 87.3 (19.2) |
| LVEF (%) | 59.6 (6.2) | 59.5 (6.2) |
| LVCO (L/min) | 5.4 (1.3) | 5.4 (1.3) |
| LVM (g) | 85.9 (22.3) | 86.2 (22.3) |
| RVEDV (mL) | 155.9 (37.1) | 156.5 (37.1) |
| RVESV (mL) | 67.3 (21.2) | 67.5 (21.3) |
| RVSV (mL) | 88.6 (20.4) | 89.0 (20.4) |
| RVEF (%) | 57.3 (6.3) | 57.3 (6.3) |
| Mean WT (mm) | 5.7 (0.8) | 5.7 (0.8) |
| Maximum WT (mm) | 9.5 (1.7) | 9.4 (1.6) |
| Mean radial strain (%) | 45.2 (8.5) | 45.2 (8.5) |
| Mean circumferential strain (%) | -22.3 (3.5) | -22.3 (3.5) |
| Binary trait | n (%) | |
| Male | 22912 (48.2) | 19509 (48.5) |
| Has ICD10 record | 40146 (84.4) | 33967 (84.5) |
| DCM | 55 (0.1) | 42 (0.1) |
| HCM | 42 (0.1) | 37 (0.1) |
| CM | 156 (0.3) | 128 (0.3) |

**Supplementary Table 2. Genome-based restricted maximum likelihood (GREML) analysis of LV spatial traits.** Table shows in the first three columns the GREML common SNPs heritability for each spatial LV trait. The next three columns shows for each spatial trait, the proportion of variance accounted by both the known confounders, including sex, age, body surface area, SBP and DBP, and the common genetic factors (through the genome relationship matrix, GRM).

| | Heritability (h2) | | | Variance explained | | |
|---|---|---|---|---|---|---|
| AHA | WT | Circ Strain | Radial Strain | WT | Circ Strain | Radial Strain |
| 1 | 19% | 17% | 14% | 46% | 21% | 18% |
| 2 | 14% | 16% | 7% | 27% | 32% | 16% |
| 3 | 16% | 16% | 15% | 32% | 18% | 27% |
| 4 | 22% | 7% | 16% | 51% | 8% | 28% |
| 5 | 29% | 11% | 15% | 58% | 18% | 21% |
| 6 | 27% | 14% | 18% | 58% | 16% | 20% |
| 7 | 25% | 14% | 16% | 64% | 24% | 24% |
| 8 | 27% | 11% | 13% | 62% | 18% | 22% |
| 9 | 27% | 24% | 17% | 67% | 28% | 26% |
| 10 | 27% | 6% | 16% | 68% | 7% | 22% |
| 11 | 30% | 11% | 16% | 67% | 16% | 23% |
| 12 | 28% | 8% | 18% | 66% | 14% | 25% |
| 13 | 16% | 17% | 14% | 53% | 24% | 21% |
| 14 | 20% | 19% | 12% | 58% | 24% | 21% |
| 15 | 15% | 21% | 18% | 53% | 23% | 24% |
| 16 | 15% | 19% | 18% | 53% | 21% | 22% |

**Supplementary Table 3. Spatial LV GWAS loci and prioritised genes.** The 42 spatial LV loci were shown by if the locus overlap with global LV loci under conventional GWS threshold, and shows which ones were reached indicated thresholds in HCM and DCM GWAS. In HCM GWAS and DCM GWAS columns, ** indicated Pval < 5e-8, * indicated FDR < 0.05. In WT, strain$^{circ}$, and strain$^{rad}$ columns, * indicates Pval < 3.125e-9 in the one of the spatial LV GWAS. Prioritised genes were mapped by positional SNPs except for the ones indicated with brackets, including those mapped by eQTL, and by chromatin interaction (CI). Full gene prioritisation table is provided in Supplementary Data Tables.

| Locus ID | CHR | Start pos (GRCh37) | Locus size (kb) | min Pval | HCM GWAS | DCM GWAS | Prioritised Gene | WT | strain$^{circ}$ | strain$^{rad}$ |
|---|---|---|---|---|---|---|---|---|---|---|
| *Spatial only* | | | | | | | | | | |
| 1 | 1 | 3197080 | 65 | 1.79E-09 | * | * | PRDM16 | | | * |
| 11 | 3 | 73544836 | 35 | 7.59E-10 | * | | PDZRN3 | | | * |
| 33 | 14 | 71697556 | 498 | 2.49E-15 | * | | SIPA1L1 | * | * | |
| 36 | 16 | 88507538 | 45 | 4.81E-11 | * | | ZFPM1 | | | * |
| 6 | 2 | 37059462 | 218 | 2.10E-12 | ** | * | STRN;HEATR5B | | * | |
| 17 | 6 | 118614518 | 413 | 1.55E-11 | ** | | CEP85L;PLN | * | | * |
| 26 | 10 | 114449904 | 67 | 1.16E-09 | ** | | VTI1A | | | * |
| 29 | 12 | 115344085 | 38 | 1.71E-11 | ** | | RNFT2,HRK(CI), TBX3 | | * | * |
| 31 | 13 | 114073950 | 5 | 1.48E-09 | ** | | ADPRHL1 | * | | |
| 15 | 5 | 132349654 | 118 | 1.90E-10 | | * | HSPA4 | * | | |
| 13 | 4 | 114380213 | 144 | 5.69E-12 | | ** | CAMK2D | * | | |
| 2 | 1 | 11827796 | 78 | 9.42E-10 | | | CLCN6 | * | | |
| 7 | 2 | 54725824 | 240 | 2.07E-11 | | | SPTBN1;EML6 | | | * |
| 14 | 5 | 64273448 | 57 | 2.06E-09 | | | CWC27 | | | * |
| 19 | 7 | 116879607 | 34 | 3.27E-10 | | | WNT2 | | * | |
| 32 | 14 | 23861811 | 12 | 7.01E-12 | | | MYH6;MYH7 | | | * |
| 38 | 17 | 37741879 | 142 | 9.39E-10 | | | ERBB2 | * | | |
| 40 | 21 | 34144545 | 39 | 1.94E-09 | | | C21orf49;C21orf62 | | | * |
| *Also in glob* | | | | | | | | | | |
| 22 | 8 | 124545147 | 7 | 7.98E-10 | * | * | FBXO32 | | * | |
| 4 | 1 | 116272483 | 60 | 6.55E-11 | * | | CASQ2 | * | | * |
| 12 | 4 | 16028096 | 9 | 1.13E-10 | * | | PROM1 | | * | |
| 20 | 8 | 8088230 | 296 | 1.91E-09 | * | | MFHAS1;ERI1 (CI) | | | * |
| 37 | 17 | 1231593 | 78 | 1.46E-10 | * | | YWHAE | * | | |
| 8 | 2 | 179381323 | 473 | 1.63E-11 | ** | * | PLEKHA3;TTN | * | * | * |
| 34 | 15 | 84488529 | 944 | 7.47E-19 | ** | * | NMB;ALPK3 | * | * | |
| 3 | 1 | 16131112 | 235 | 5.14E-12 | ** | ** | HSPB7;CLCNKA | | * | * |
| 16 | 6 | 36618821 | 41 | 2.60E-17 | ** | ** | CDKN1A | * | * | * |
| 23 | 8 | 125849614 | 52 | 2.90E-10 | ** | ** | MTSS1;SQLE (eQTL) | | * | |
| 27 | 10 | 121414236 | 34 | 3.64E-15 | ** | ** | BAG3 | | * | |
| 39 | 17 | 43463493 | 1402 | 1.32E-15 | ** | ** | MAPT;KANSL1;WNT3 | * | | |
| 41 | 22 | 24111044 | 71 | 9.78E-14 | ** | ** | SMARCB1;DERL3 | * | * | |
| 24 | 10 | 75404300 | 181 | 5.05E-11 | ** | | MYOZ1;SYNPO2L | * | | |
| 28 | 11 | 47365014 | 632 | 9.73E-10 | ** | | MYBPC3;SPI1 | * | | |
| 5 | 1 | 236841577 | 13 | 1.11E-12 | | ** | ACTN2 | | * | |
| 9 | 2 | 218251702 | 62 | 8.29E-10 | | | DIRC3 | * | | |
| 10 | 3 | 69796492 | 113 | 2.54E-12 | | | MITF | | | * |
| 18 | 7 | 46609344 | 59 | 7.95E-10 | | | IGFBP1;IGFBP3(CI) | * | | |
| 21 | 8 | 11776904 | 60 | 9.92E-13 | | | DEFB136;DEFB135 | | * | * |
| 25 | 10 | 112544125 | 0 | 5.27E-10 | | | RBM20 | | * | |
| 30 | 12 | 120646830 | 52 | 1.41E-09 | | | PXN | * | | |
| 35 | 15 | 99249029 | 47 | 2.72E-15 | | | IGF1R | * | | |
| 42 | 22 | 26155484 | 9 | 8.34E-19 | | | MYO18B | * | | * |

**Supplementary Table 4. Cross check Spatial GWAS genes with Cardiac G2P database.** Cardiac G2P mapped genes were listed by disease grouping[18].

| Cardiac Disease grouping | Cardiac G2P mapped genes | Found in Spatial LV GWAS |
|---|---|---|
| Classic CPVT phenotype | RYR2,CASQ2 | CASQ2 |
| Familial dilated cardiomyopathy | BAG3,DES,DSP,FLNC,LMNA, MYH7,PLN,RBM20,SCN5A, TNNC1,TNNT2, TTN | BAG3,MYH7, PLN,RBM20, TTN |
| Familial hypertrophic cardiomyopathy | ACTC1,MYBPC3,MYH7,MYL2, MYL3,PLN,TNNI3,TNNT2, TPM1 | MYBPC3,MYH7, PLN |
| Rare syndromic disorder with hypertrophic cardiomyopathy - isolated LVH | ALPK3,CACNA1C,DES,FHL1, FLNC,GLA,LAMP2, PRKAG2,PTPN11, PTPN11,RAF1,RIT1,TTR | ALPK3 |
| Syndrome with hypertrophic cardiomyopathy - no isolated LVH | ABCC9,BAG3,CRYAB, FXN,GAA,MYO6,SLC25A4 | BAG3 |

**Supplementary Table 5. The Exome variants significantly associated (min *P* < 3.125e-9) with spatial LV traits, but were not included in the impute data on which GWAS were performed.** The list shows 19 Exome variants, and for each variant its location in GRCh38, the effect allele (EA), the effect allele frequency (EAF), the most severe Ensembl consequence (VEP), and the gene symbols.

| GWAS locus ID | CHR | POS | EA | NEA | EAF | VEP | Gene(s) |
|---|---|---|---|---|---|---|---|
| 6 | 2 | 36883902 | A | T | 0.560971 | intron | STRN |
| 6 | 2 | 37007376 | C | T | 0.581296 | intron | HEATR5B |
| 8 | 2 | 178612605 | A | AT | 0.212555 | intron | AC010680.4, TTN, TTN-AS1 |
| 16 | 6 | 36678991 | CTA | C | 0.195241 | intron | CDKN1A, DINOL, LAP3P2, PANDAR |
| 16 | 6 | 36679011 | G | A | 0.326233 | intron | CDKN1A, DINOL, LAP3P2, PANDAR |
| - | 10 | 43616994 | G | A | 7.80E-05 | synonymous | ZNF485 |
| 24 | 10 | 73650119 | T | C | 0.149312 | intron | AC073389.2, SYNPO2L |
| 24 | 10 | 73682785 | T | C | 0.136494 | missense | AGAP5, BMS1P4-AGAP5 |
| - | 11 | 19192439 | G | A | 0.00497613 | missense | CSRP3, CSRP3-AS1 |
| 39 | 17 | 45816403 | G | GC | 0.222562 | intron | CRHR1, LINC02210-CRHR1, MAPT-AS1 |
| 39 | 17 | 45816406 | T | TGCCTG | 0.222557 | intron | CRHR1, LINC02210-CRHR1, MAPT-AS1 |
| 39 | 17 | 46171448 | A | G | 0.223671 | synonymous | KANSL1 |
| 39 | 17 | 46171471 | C | T | 0.186527 | missense | KANSL1 |
| 39 | 17 | 46171730 | C | A | 0.1757 | synonymous | KANSL1 |
| 39 | 17 | 46171833 | G | T | 0.172555 | missense | KANSL1 |
| 39 | 17 | 46548981 | C | T | 0.198277 | missense | ARL17A, LRRC37A2 |
| 39 | 17 | 46704913 | G | C | 0.179936 | intron | NSF |
| - | 17 | 64897294 | G | A | 0.195414 | 5 prime UTR | AC103810.2, AC103810.5, AC103810.7, LRRC37A3 |
| 42 | 22 | 25768112 | T | C | 0.433029 | splice region | MYO18B |

**Supplementary Table 6. List of Exome loss-of-function or splice variants observed in the GWAS loci.** Genes and consequence came from running VEP, 'genes' column included all gene symbols that the variant might influence, and 'consequence' column shows the most severe consequence. Beta (max) and MLOG1OP (max) were respectively the largest absolute beta values and the largest -log10 of P values from testing the 16 segments of LV on WT, radial and circumferential strain, the LVEF, LVM and global max and mean WT, global mean radial and circumferential strain.

| GenomicLocus | Gene(s) | Variant (GRCh38:NEA:EA) | MAF | Consequence | BETA (max) | MLOG10P (max) |
|---|---|---|---|---|---|---|
| 2 | C1orf167 | chr1:11768081:C:T | 0.00017152 | stop gained | 0.773755 | 2.03526 |
| 2 | C1orf167 | chr1:11768102:C:T | 7.80E-05 | stop gained | -0.487175 | 0.858324 |
| 2 | C1orf167 | chr1:11775536:G:A | 7.80E-05 | stop gained | 0.993422 | 1.70652 |
| 2 | C1orf167 | chr1:11784298:C:T | 0.00062369 | stop gained | 0.432227 | 2.33912 |
| 2 | C1orf167 | chr1:11784463:C:T | 0.0288459 | stop gained | -0.0583737 | 2.00594 |
| 2 | C1orf167 | chr1:11785177:TGC:T | 7.80E-05 | frameshift variant | 0.639418 | 0.87988 |
| 2 | C1orf167 | chr1:11789438:C:T | 0.00012474 | stop gained | 0.731767 | 1.95578 |
| 3 | CLCNKA | chr1:16032247:C:T | 0.00046777 | stop gained | -0.401888 | 1.71203 |
| 3 | CLCNKA | chr1:16026765:TCCCTTCAGCGGTGAGACCCCCCTCATGCCCCGCC:T | 0.00065691 | splice donor variant | -0.409451 | 2.36107 |
| 3 | CLCNKA | chr1:16028061:C:T | 0.00038981 | stop gained | 0.388967 | 2.44515 |
| 3 | ZBTB17 | chr1:15946317:C:G | 7.80E-05 | splice acceptor variant | -1.01736 | 1.80327 |
| 8 | TTN | chr2:178579702:G:A | 7.80E-05 | stop gained | -1.43759 | 3.1917 |
| 8 | TTN-AS1 | chr2:178615321:A:G | 0.00029626 | splice acceptor variant | 0.414412 | 2.12027 |
| 8 | CCDC141 | chr2:178837693:G:A | 0.0004054 | stop gained | -0.318306 | 1.17162 |
| 8 | CCDC141 | chr2:178978618:TC:T | 0.00054573 | frameshift variant | -0.371958 | 1.86696 |
| 8 | CCDC141 | chr2:178834173:T:A | 0.00035862 | stop lost | -0.341691 | 1.3302 |
| 8 | TTN | chr2:178528273:C:T | 7.80E-05 | splice donor variant | -0.842612 | 1.46746 |
| 8 | TTN | chr2:178653473:CT:C | 0.0002027 | frameshift variant | -0.446397 | 1.06941 |
| 8 | TTN | chr2:178662420:T:A | 9.36E-05 | splice acceptor variant | 0.627603 | 1.38918 |
| 8 | TTN | chr2:178663903:C:T | 7.80E-05 | splice acceptor variant | -1.0964 | 2.76886 |
| 8 | TTN | chr2:178664443:GACAGTTAAGAATGTACCTTTGACAGGTACA:G | 0.00032907 | splice donor variant | 0.407317 | 1.36414 |
| 8 | TTN | chr2:178665777:G:A | 0.00014034 | stop gained | -0.746017 | 1.75502 |
| 8 | TTN | chr2:178677634:TG:T | 0.00015592 | frameshift variant | -0.732263 | 2.67767 |
| 8 | TTN | chr2:178689289:C:A | 0.00010915 | splice donor variant | -0.770963 | 1.45558 |
| 8 | TTN | chr2:178689897:C:T | 0.00059257 | splice acceptor variant | -0.238994 | 0.938127 |
| 8 | TTN | chr2:178745884:C:A | 0.00014033 | stop gained | 0.913398 | 2.35187 |
| 8 | TTN | chr2:178746047:G:C | 9.36E-05 | stop gained | 0.730598 | 1.23884 |
| 8 | TTN | chr2:178749346:CCCTG:C | 0.00012474 | frameshift variant | 0.808616 | 1.71903 |
| 8 | TTN | chr2:178749351:ATG:A | 0.00012474 | frameshift variant | 0.808616 | 1.71903 |
| 8 | TTN | chr2:178749358:TGC:T | 0.00012474 | frameshift variant | 0.808616 | 1.71903 |
| 8 | TTN | chr2:178751617:AT:A | 0.00010915 | frameshift variant | 0.814172 | 1.60621 |
| 8 | TTN | chr2:178756292:T:TC | 0.00049897 | frameshift variant | 0.35086 | 2.07556 |
| 8 | TTN | chr2:178758982:A:G | 7.80E-05 | splice donor variant | -1.1297 | 2.27518 |
| 8 | TTN-AS1 | chr2:178778004:G:A | 7.80E-05 | splice donor variant | -1.46599 | 5.08606 |
| 17 | CEP85L | chr6:118465397:ACACTTGAT:A | 9.36E-05 | stop lost | 1.04589 | 2.19619 |
| 17 | CEP85L | chr6:118652721:CTGAT:C | 0.00037445 | frameshift variant | -0.406472 | 1.46053 |
| 20 | PRAG1 | chr8:8378000:G:A | 0.00023389 | stop gained | 0.655165 | 2.19556 |
| 20 | PRAG1 | chr8:8378066:G:A | 0.00010915 | stop gained | -0.813455 | 2.41299 |
| 24 | NDST2 | chr10:73808145:G:A | 7.80E-05 | stop gained | 1.05322 | 2.18563 |
| 24 | FUT11 | chr10:73772572:G:A | 9.36E-05 | stop gained | -1.14761 | 2.39067 |
| 24 | FUT11 | chr10:73773484:G:T | 0.00018711 | stop gained | -0.882416 | 2.75779 |
| 24 | AGAP5 | chr10:73674703:G:A | 0.00018711 | stop gained | 0.639122 | 1.63075 |
| 24 | AGAP5 | chr10:73675604:A:T | 9.36E-05 | stop gained | -0.773698 | 1.8186 |
| 24 | SYNPO2L | chr10:73653643:C:A | 0.00014033 | stop gained | 0.84727 | 2.02775 |
| 24 | AGAP5 | chr10:73682707:T:TTG | 9.36E-05 | frameshift variant | -0.745795 | 1.86109 |
| 24 | SYNPO2L | chr10:73647531:A:AG | 0.00086754 | frameshift variant | 0.378353 | 2.39022 |
| 28 | AGBL2 | chr11:47667666:TC:T | 0.00014034 | frameshift variant | 0.792264 | 2.15626 |
| 28 | C1QTNF4 | chr11:47590761:AGGGCCCAGCAGGCCGCT:A | 0.00042143 | frameshift variant | -0.434965 | 1.75612 |
| 28 | C1QTNF4 | chr11:47590762:G:GGGCCCAGCAGGCCGCTGGGCCCAGCA | 0.00015608 | frameshift variant | -0.727352 | 1.91261 |
| 28 | C1QTNF4 | chr11:47590762:GGGCCCAGCAGGCCGCTGGGCCCAGCA:G | 0.00012487 | frameshift variant | 0.698676 | 1.36325 |
| 28 | FAM180B | chr11:47588126:C:CT | 0.00042099 | frameshift variant | -0.356831 | 1.35924 |
| 28 | FAM180B | chr11:47588313:G:A | 0.0002027 | stop gained | 0.577382 | 1.5162 |
| 28 | KBTBD4 | chr11:47573330:T:TC | 7.80E-05 | frameshift variant | 0.739412 | 1.14225 |
| 28 | KBTBD4 | chr11:47578911:C:G | 9.36E-05 | splice donor variant | -1.0408 | 2.11123 |
| 28 | NDUFS3 | chr11:47579334:CGTGA:C | 0.00023389 | splice donor variant | -0.476536 | 1.35272 |
| 28 | NDUFS3 | chr11:47580713:TA:T | 7.80E-05 | frameshift variant | -0.810483 | 1.83795 |
| 28 | PTPMT1 | chr11:47571628:G:C | 0.00394487 | stop lost | -0.0934814 | 0.929256 |
| 28 | SLC39A13 | chr11:47415038:G:A | 9.36E-05 | splice acceptor variant | 0.694338 | 1.10392 |
| 28 | RAPSN | chr11:47438712:AC:A | 9.39E-05 | frameshift variant | 1.25403 | 2.85113 |
| 30 | PXN | chr12:120216728:AC:A | 0.00032744 | frameshift variant | 0.339037 | 0.95313 |
| 31 | ADPRHL1 | chr13:113424310:G:A | 7.80E-05 | stop gained | 1.05364 | 1.80184 |
| 34 | ALPK3 | chr15:84856391:G:A | 9.36E-05 | splice acceptor variant | 1.2655 | 3.51923 |
| 34 | ADAMTSL3 | chr15:83838216:G:A | 0.00014034 | splice donor variant | -0.55498 | 1.31167 |
| 34 | ALPK3 | chr15:84817508:CG:C | 0.00021758 | frameshift variant | -0.709746 | 2.06313 |
| 34 | ALPK3 | chr15:84863641:G:GT | 0.00010915 | splice donor variant | 1.04933 | 2.77413 |
| 34 | ALPK3 | chr15:84868257:C:CT | 0.00014033 | frameshift variant | -0.757259 | 1.78261 |
| 34 | WDR73 | chr15:84643474:CG:C | 0.00098235 | frameshift variant | 0.237993 | 1.29681 |
| 34 | WDR73 | chr15:84643679:C:A | 9.36E-05 | stop gained | -0.75819 | 1.67919 |
| 34 | WDR73 | chr15:84654233:C:G | 0.00012474 | splice donor variant | 0.765469 | 1.64125 |
| 34 | SLC28A1 | chr15:84887782:C:T | 0.00028066 | stop gained | 0.414566 | 1.3446 |
| 38 | PGAP3 | chr17:39672852:T:C | 0.00017152 | stop lost | 0.591775 | 1.42737 |
| 38 | PNMT | chr17:39670186:GC:G | 0.00020271 | frameshift variant | 0.585403 | 1.61095 |
| 38 | STARD3 | chr17:39660527:G:A | 9.36E-05 | splice donor variant | 0.890304 | 1.56795 |
| 39 | LRRC37A2 | chr17:46548749:A:T | 0.00012474 | stop gained | 0.844679 | 1.88409 |
| 39 | LRRC37A2 | chr17:46548914:TC:T | 7.80E-05 | frameshift variant | -0.907647 | 1.49185 |
| 39 | KANSL1 | chr17:46094507:C:T | 0.0003119 | splice donor variant | -0.524346 | 1.82778 |
| 39 | MAPT | chr17:45983226:G:GC | 0.00014033 | frameshift variant | -0.599284 | 1.32848 |
| 39 | MAPT | chr17:45985745:G:A | 0.00010915 | splice donor variant | -0.758089 | 1.45605 |
| 40 | C21orf62 | chr21:32794101:G:T | 0.00023389 | stop gained | -0.421833 | 1.06357 |
| 41 | DERL3 | chr22:23838364:GC:G | 0.00163725 | frameshift variant | -0.241464 | 1.99186 |
| 41 | MMP11 | chr22:23772980:T:C | 7.80E-05 | splice donor variant | 0.906691 | 1.50627 |

### Supplementary Figures

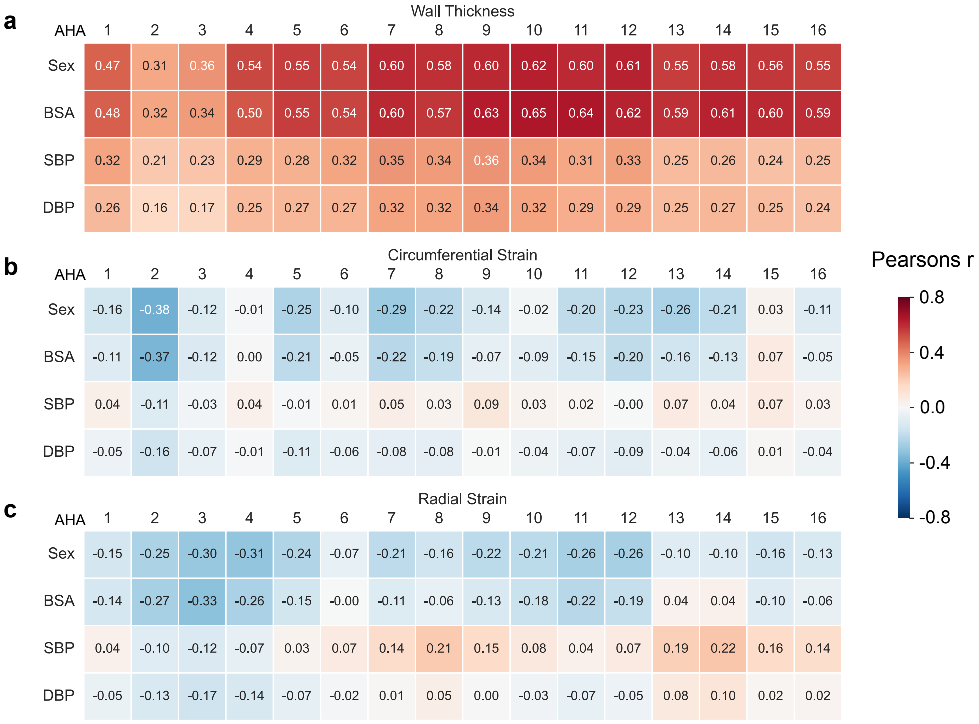


**Supplementary Figure 1. LV spatial traits correlations to known confounding factors.** Association of the spatial wall thickness (a), strain$^{circ}$ (b) and strain$^{rad}$ (c) with Sex, body surface area (BSA), systolic (SBP) and diastolic (DBP) blood pressure. Pearson correlation values are shown for each indicated regional trait and the indicated known predictor on the left. Values were calculated with DataFrame.corr function in pandas v2.1.3.

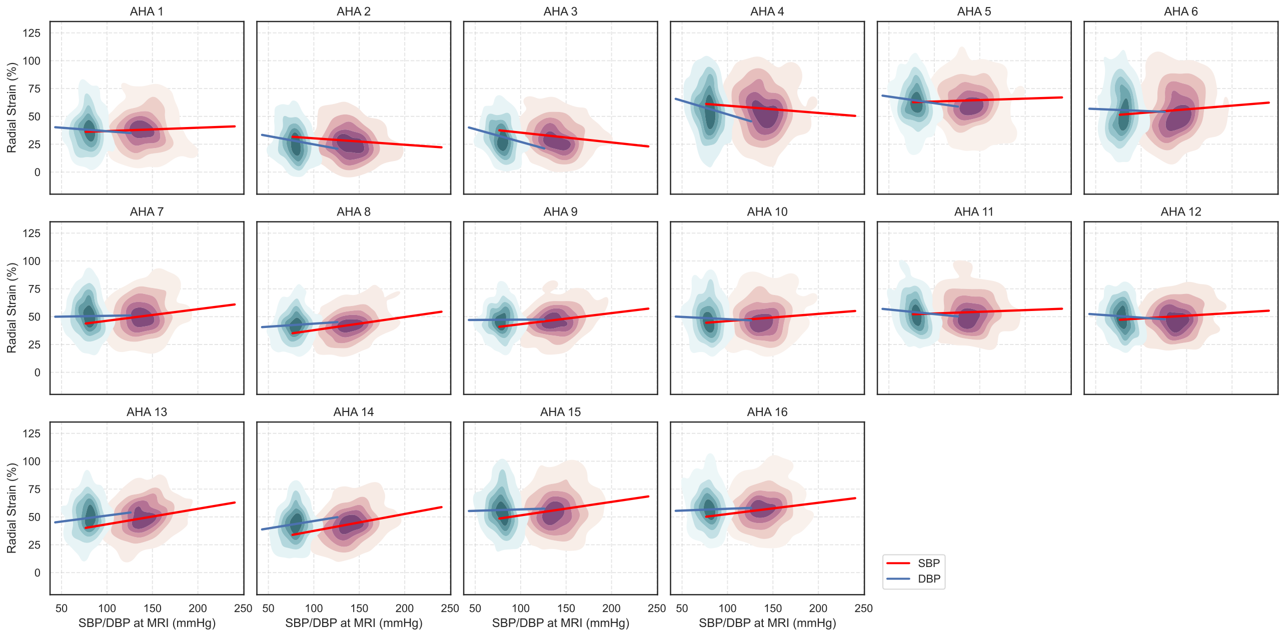


**Supplementary Figure 2. Association of LV spatial strain$^{rad}$ with systolic (SBP) and diastolic (DBP) blood pressure.** Each graph displays a kernel density plot of mean strain$^{rad}$ on an AHA segment against automatic SBP (red cloud) and DBP (blue cloud) readings taken at MRI. $n = 40,058$ participants were included in the analysis. Plot produced with kdeplot and regplot functions in seaborn v0.12.2.

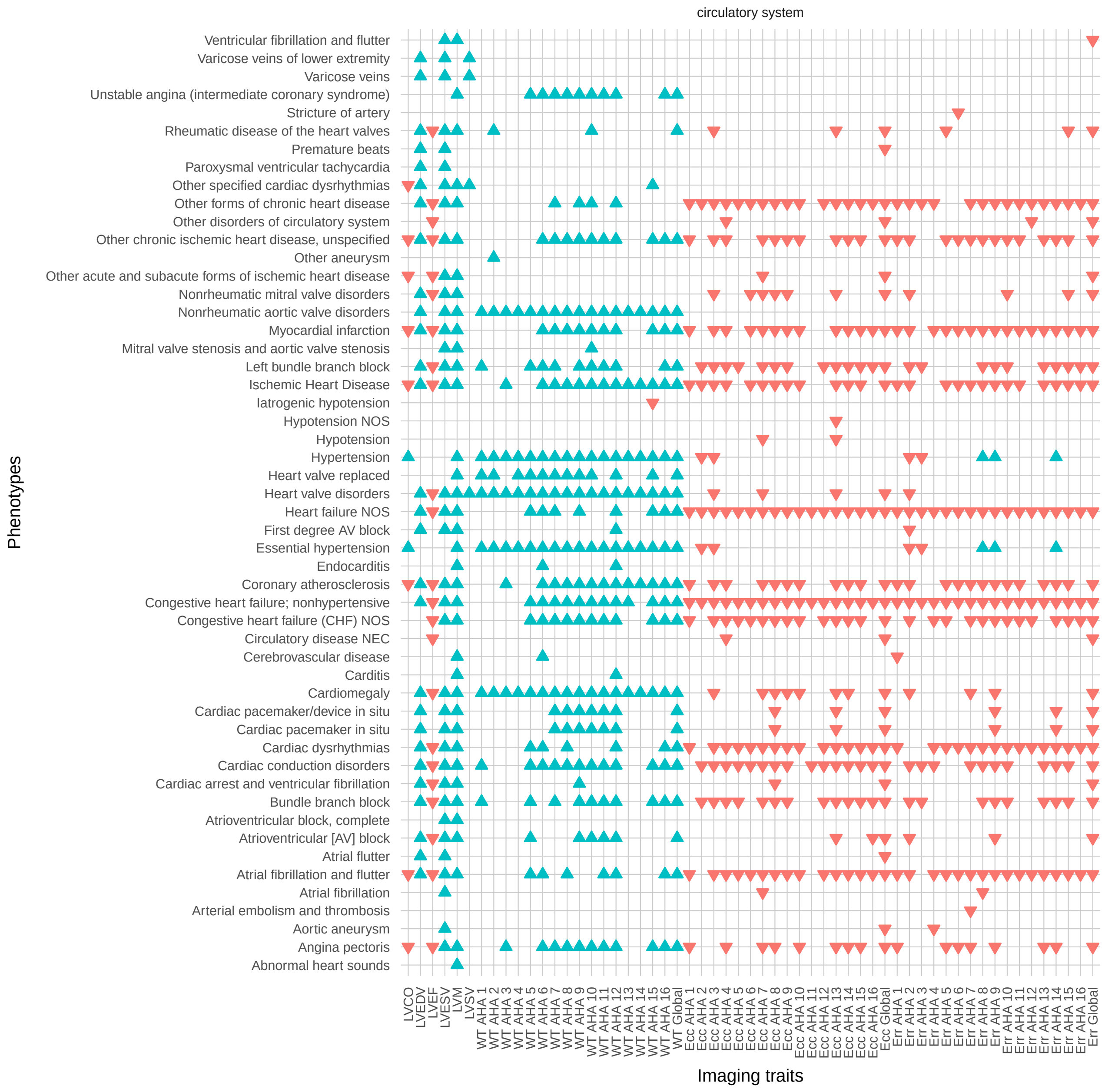


**Supplementary Figure 3. Phenome-wide association study of spatial left ventricular traits on cardiovascular phenotypes.** After adjustment for known confounders (sex, age, body surface area, SBP, DBP), each spatial trait was assessed for association with cardiovascular phenotypes. Phenotypes as phecodes are described on the y-axis with the phecode category separating the groups and the imaging traits are on the x-axis. Each point denotes a significant PheWAS association with a Bonferroni correction for 1,840 analyzed phecodes. The shape and colour denote the direction of effect. See the Supplementary Data for the full PheWAS results.

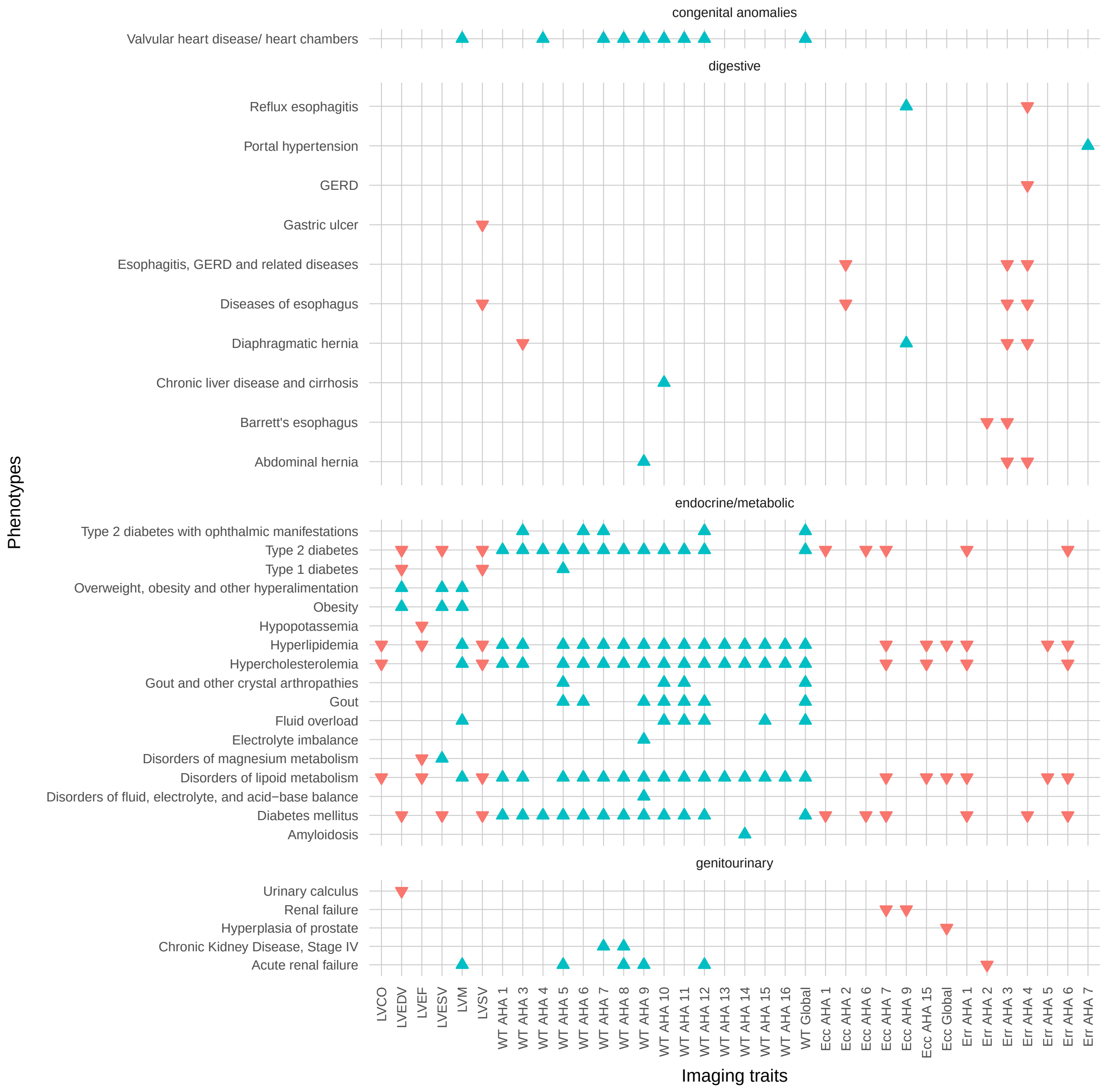


**Supplementary Figure 4. Phenome-wide association study of spatial left ventricular traits on non-cardiovascular phenotypes.** After adjustment of known confounders (sex, age, body surface area, SBP, DBP), each spatial trait was assessed for association with non-cardiovascular phenotypes. Phenotypes as phecodes are described on the y-axis with the phecode category separating the groups and the imaging traits are on the x-axis. Each point denotes a significant PheWAS association with a Bonferroni correction for 1,840 analyzed phecodes. The shape and colour denote the direction of effect. See the Supplementary Data for the full PheWAS results.

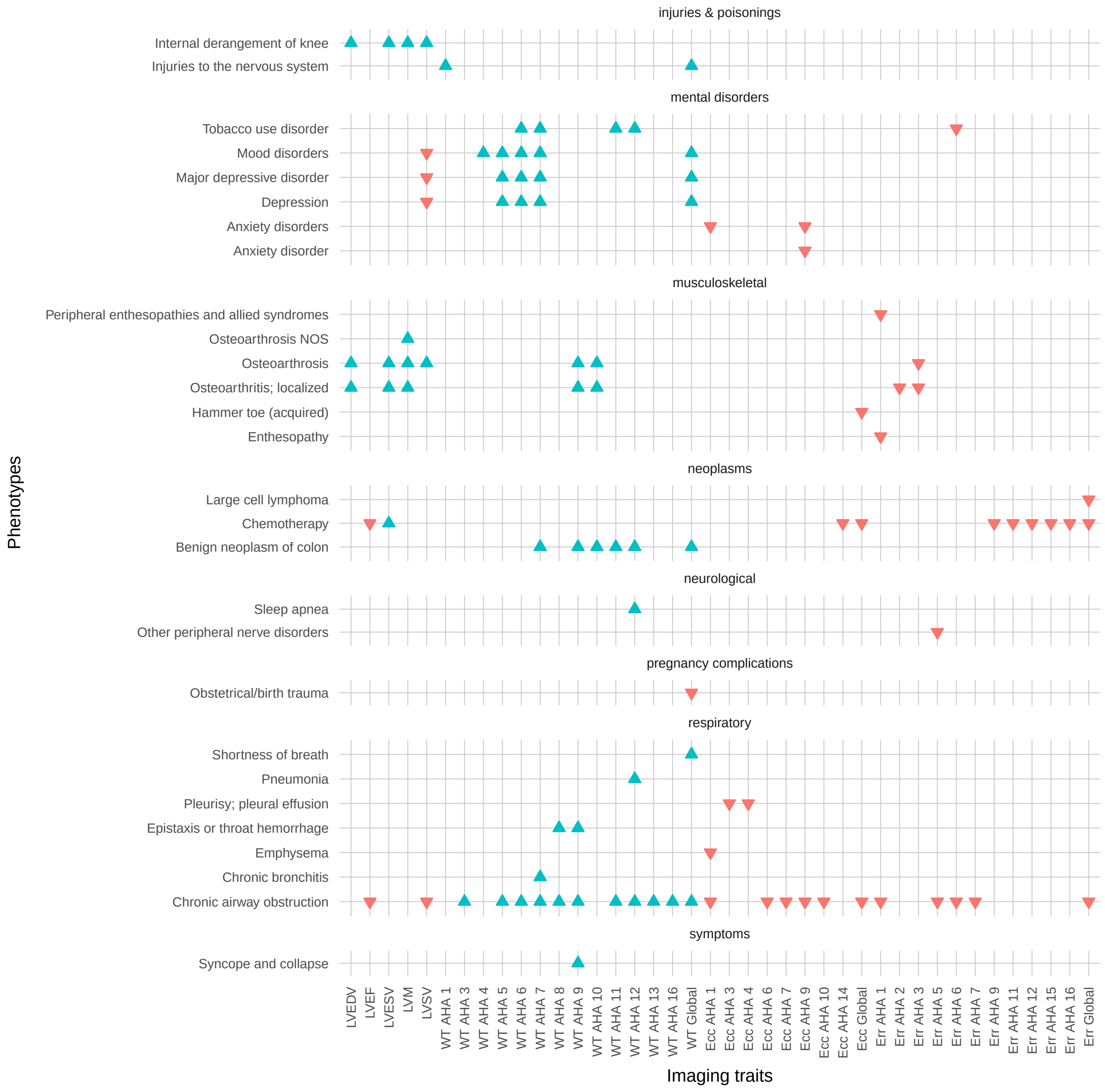


**Supplementary Figure 5. Phenome-wide association study of spatial left ventricular traits on additional non-cardiovascular phenotypes.** After adjustment of known confounders (sex, age, body surface area, SBP, DBP), each spatial trait was assessed for association with non-cardiovascular phenotypes. Phenotypes as phecodes are described on the y-axis with the phecode category separating the groups and the imaging traits are on the x-axis. Each point denotes a significant PheWAS association with a Bonferroni correction for 1,840 analyzed phecodes. The shape and colour denote the direction of effect. See the Supplementary Data for the full PheWAS results.

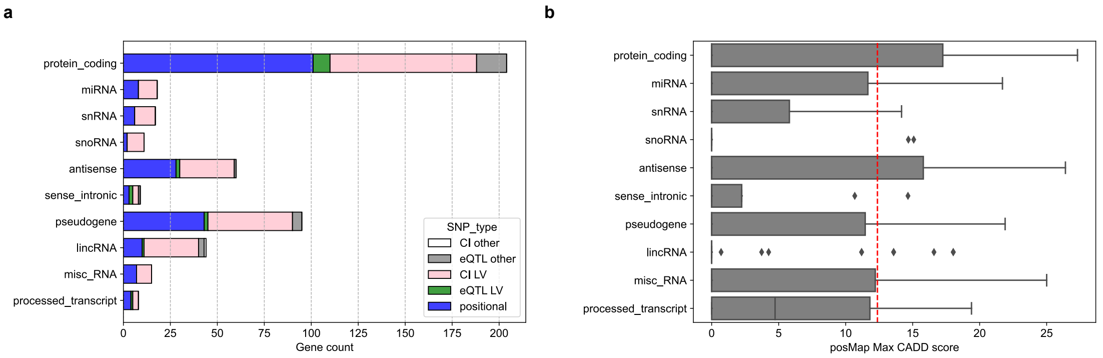


**Supplementary Figure 6. Spatial GWAS gene types by FUMA**. (a) Distribution of gene products identified in the 42 loci by gene types, prioritised by: (1) if significant SNPs were found in the gene product (blue), (2) if the gene product contains no such positional SNPs, gene products that were mapped by cis-eQTL variants in the heart left ventricle (GTEx8) (green), (3) if none above were found, if they were mapped by chromatin interaction (CI) overlap in left ventricle (pink), and lastly, if they are mapped by eQTL and CI in other 4 tissues (Heart Aorta Appendage, Arteria, Artery A, and Atery Tibial). Total number of gene products was 482, and 455 (94%) were mapped by positional genes, or eQTL or CI in heart left ventricle. (b) Distribution of maximum CADD scores for gene products where positional SNPs were found. The gene products and CADD scores were annotated using FUMA. Legends. miRNA: small RNA (22bp) that silences the expression of target mRNA; snRNA: Small RNA in the cell nucleus involved in the processing of pre messenger RNAs; snoRNA: Small RNA in the cell nucleolus involved in the post-transcriptional modification of other RNAs, antisense: transcripts on the opposite strand, sense intronic: long non-coding transcript in introns of a coding gene, pseudogene: a gene that has homology to known protein-coding genes but contain a frameshift and/or stop codon(s) which disrupts the open reading frame (ORF). Thought to have arisen through duplication followed by loss of function, lincRNA: long intergenic ncRNA, misc RNA: miscellaneous RNA that cannot be classified, processed transcript: transcript that doesn't contain an ORF.

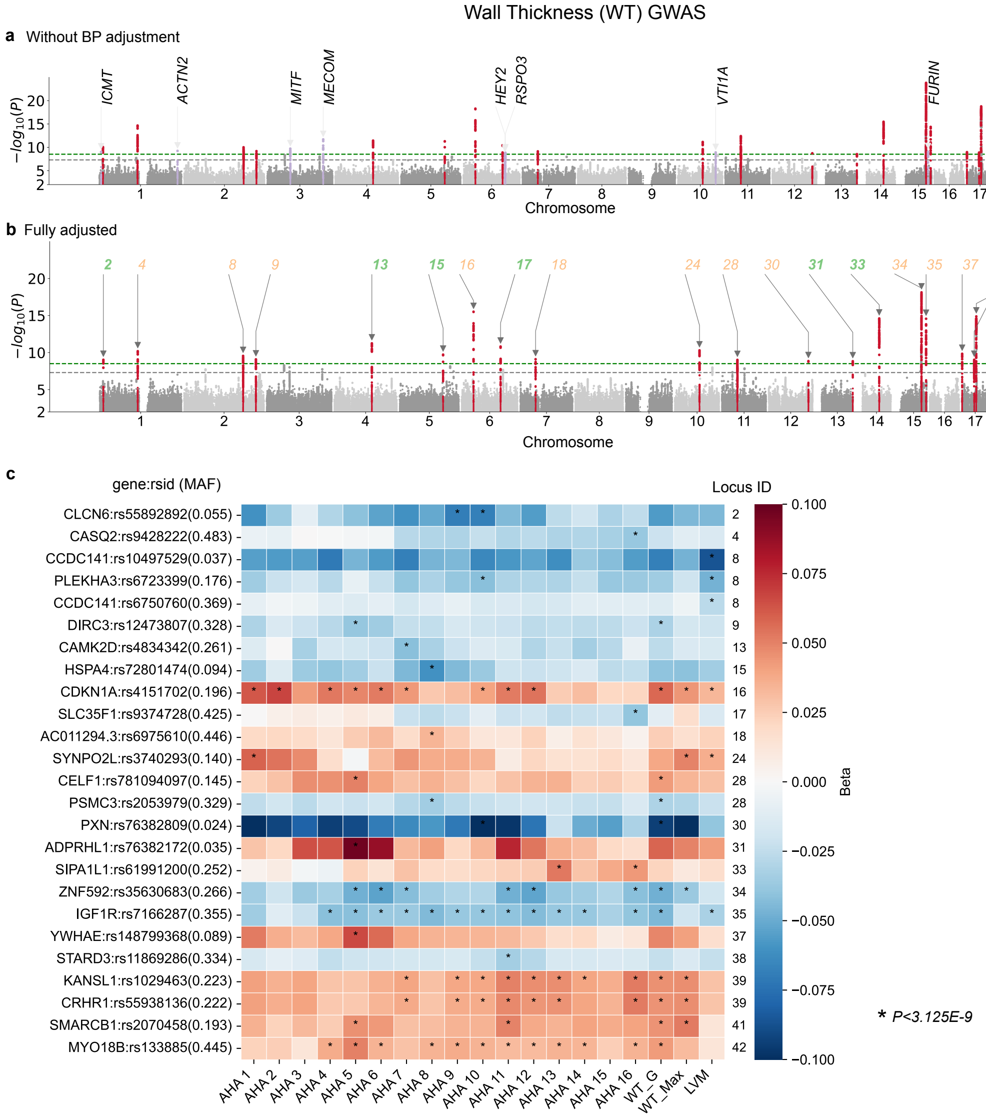


**Supplementary Figure 7. Manhattan plots for the wall thickness spatially resolved by the AHA-16 model.** (a) The wall thicknesses is adjusted by Sex, age at MRI, age squared, BMI, BSA and the top 10 PCs. Minimum P value found by GWAS on 16 segments were shown in the manhattan plot. Highlights in red show the loci identified in fully adjusted GWAS (panel b), highlights in purple show additional loci that were no longer significant after adjustment with BP. These loci (purple) were annotated by the nearest gene. (b) The wall thicknesses are additionally adjusted by SBP and DBP measured at MRI. (c) Heatmap showing the beta values of lead SNPs in each locus (by fully adjusted GWAS) with the AHA segments on LV.

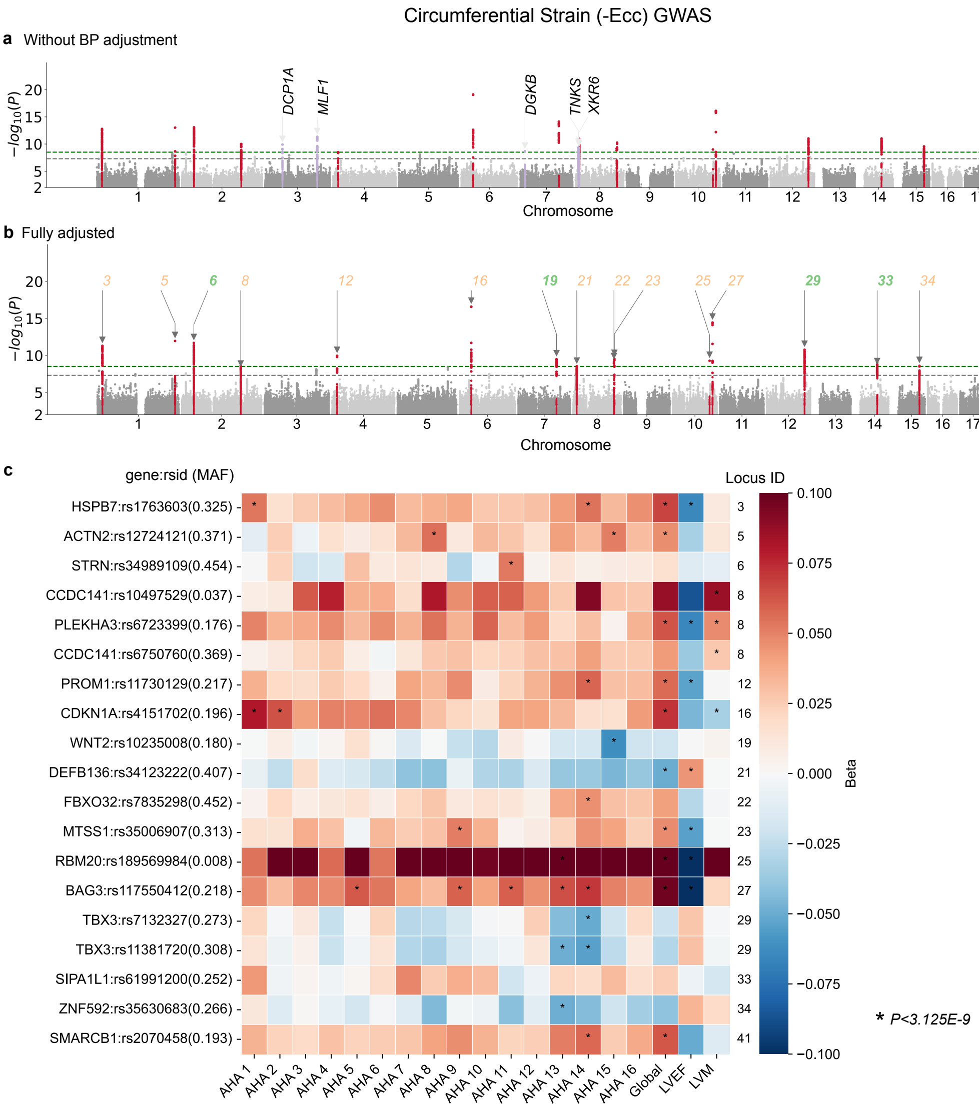


**Supplementary Figure 8. Manhattan plots for the circumferential strain spatially resolved by the AHA-16 model.** (a) The circumferential strain is adjusted by Sex, age at MRI, age squared, BMI, BSA and the top 10 PCs. Minimum P value found by GWAS on 16 segments were shown in the manhattan plot. Highlights in red show the loci identified in fully adjusted GWAS (panel b), highlights in purple show additional loci that were no longer significant after adjustment with BP. These loci (purple) were annotated by the nearest gene. (b) The circumferential strain are additionally adjusted by SBP and DBP measured at MRI. (c) Heatmap showing the beta values of lead SNPs in each locus (by fully adjusted GWAS) with the AHA segments on LV.

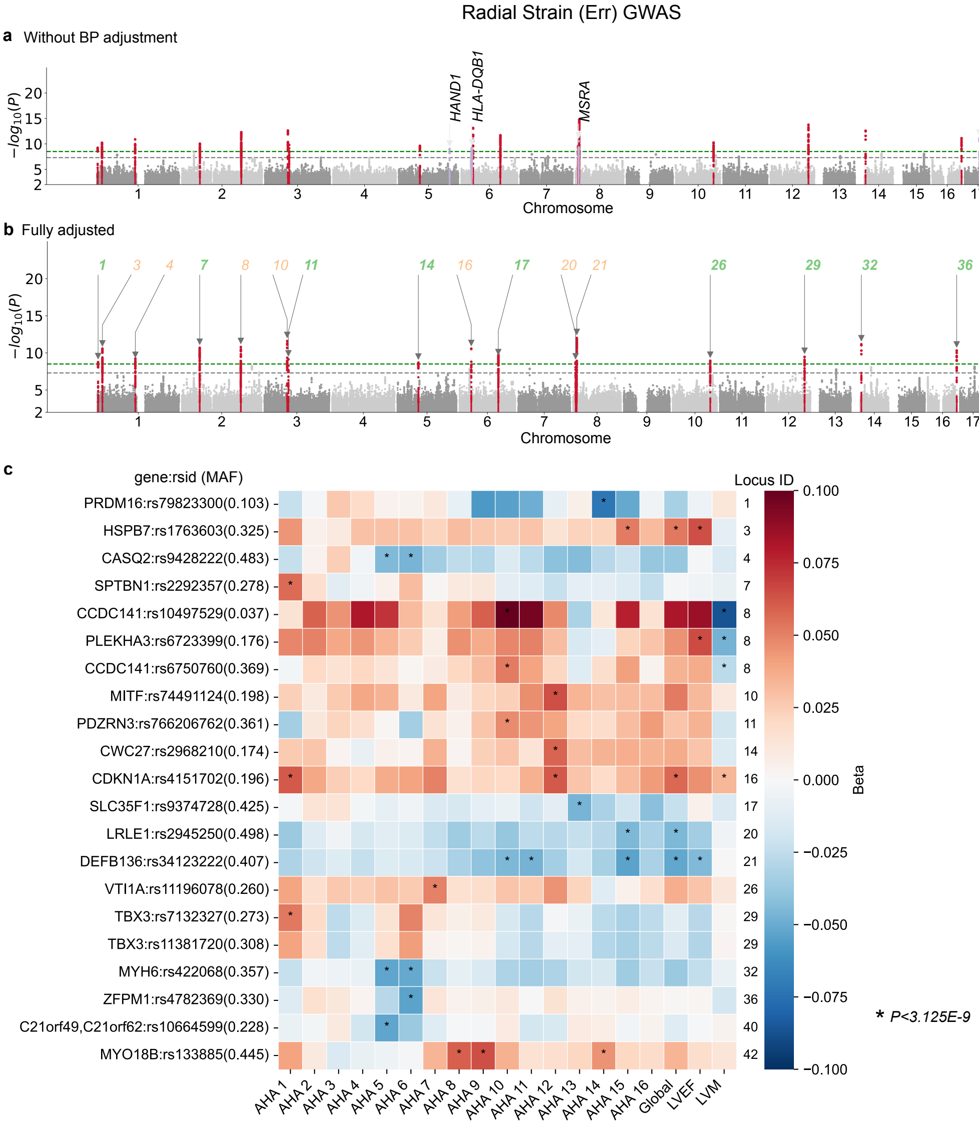


**Supplementary Figure 9. Manhattan plots for the radial strain spatially resolved by the AHA-16 model.**. (a) The radial strain is adjusted by Sex, age at MRI, age squared, BMI, BSA and the top 10 PCs. Minimum P value found by GWAS on 16 segments were shown in the manhattan plot. Highlights in red show the loci identified in fully adjusted GWAS (panel b), highlights in purple show additional loci that were no longer significant after adjustment with BP. These loci (purple) were annotated by the nearest gene. (b) The radial strain are additionally adjusted by SBP and DBP measured at MRI. (c) Heatmap showing the beta values of lead SNPs in each locus (by fully adjusted GWAS) with the AHA segments on LV.

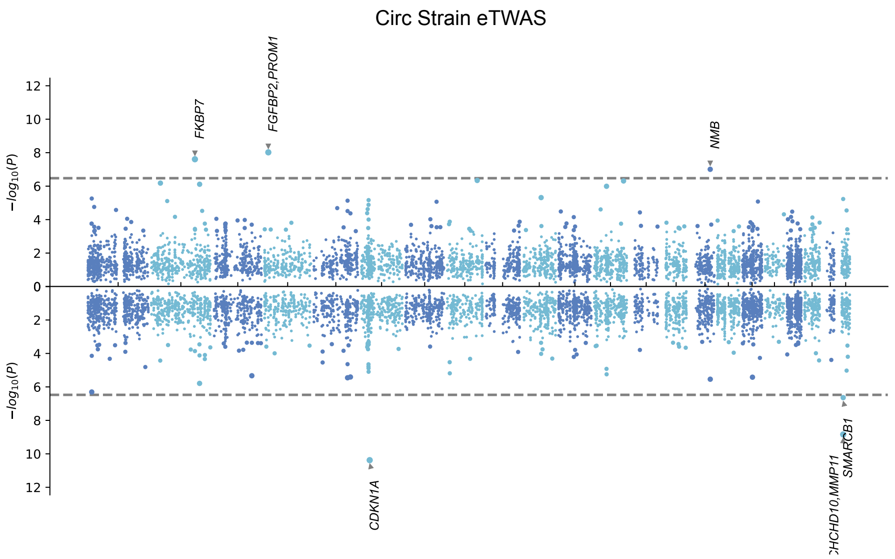


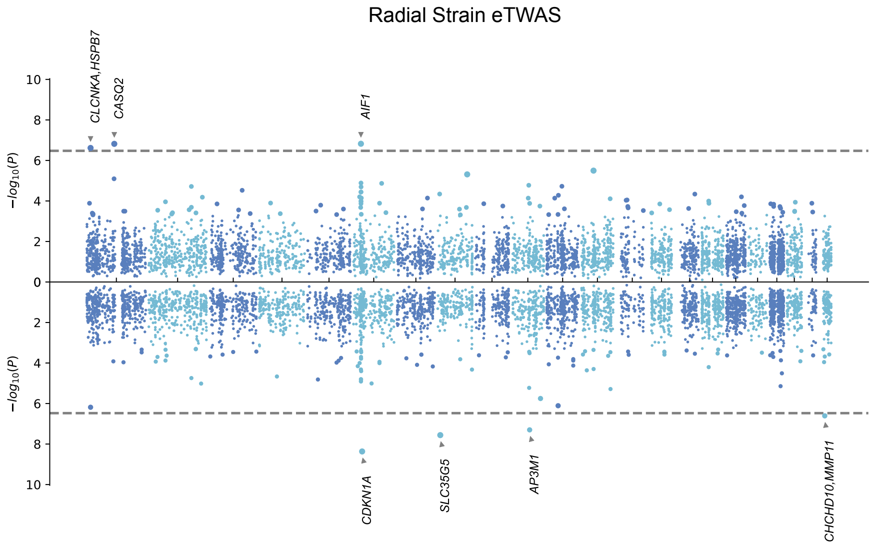


**Supplementary Figure 10. Predicted regulatory effects of GWAS variants on expression.** Regulatory effects were calculated using GWAS summary statistics and GTEx v8 eQTL MASH-R model for the heart left ventricle. 10,498 genes were tested. Manhattan plot shows the minimum P-value on the 16 regional traits of strain$^{circ}$ and strain$^{rad}$. Related to Figure 5.

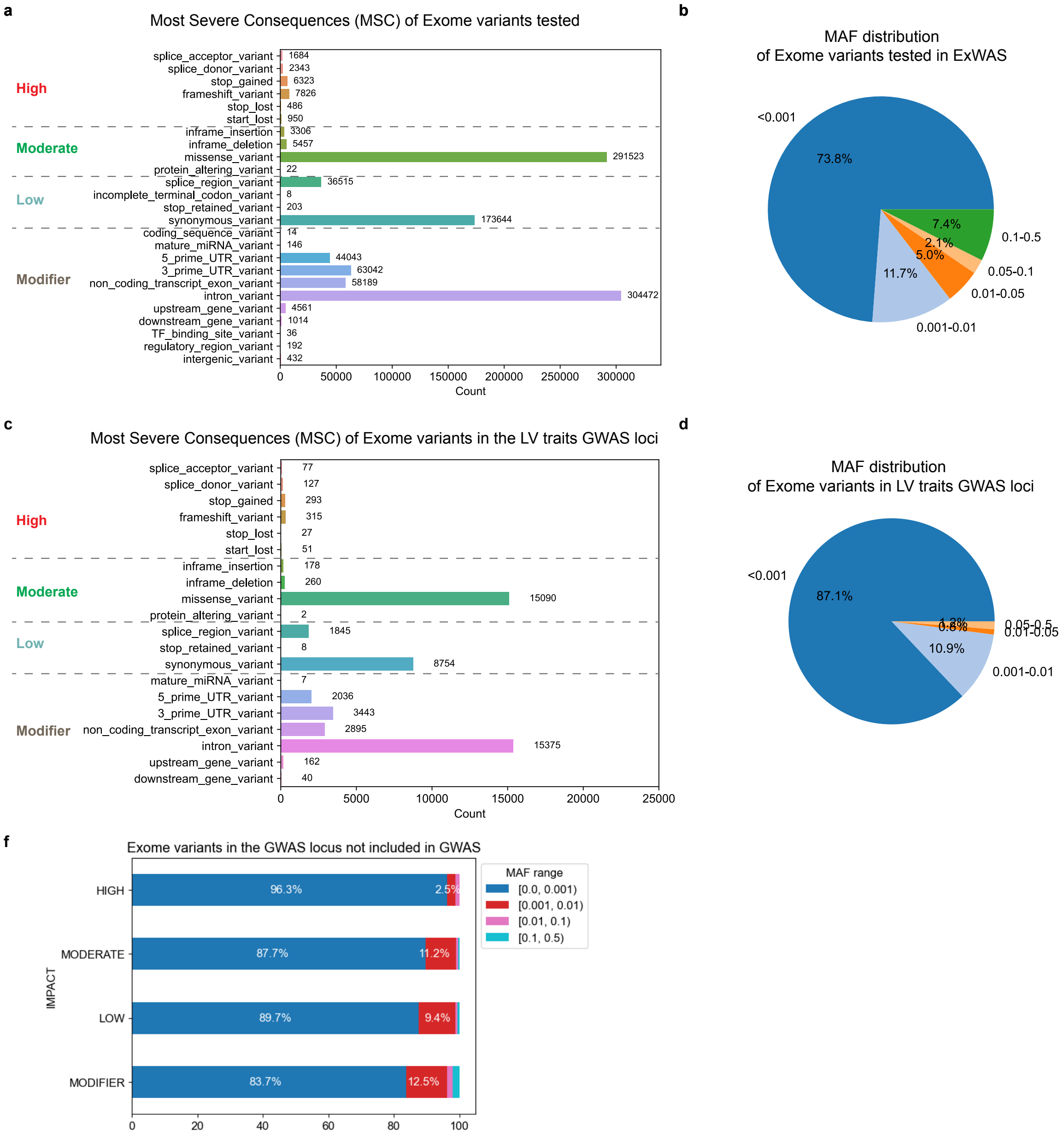


**Supplementary Figure 11. Variant effect annotations of whole Exome data in the UKBB CMR imaging cohort.** (a) Number of variants tested in ExWAS by the most severe consequence (MSC) annotated with Ensembl Variant Effect Predictor (VEP). 1,006,431 Exome variants that appeared at least five times in the UKBB CMR cohort were tested. (b) Distribution of the variants tested by minor allele frequency (MAF).

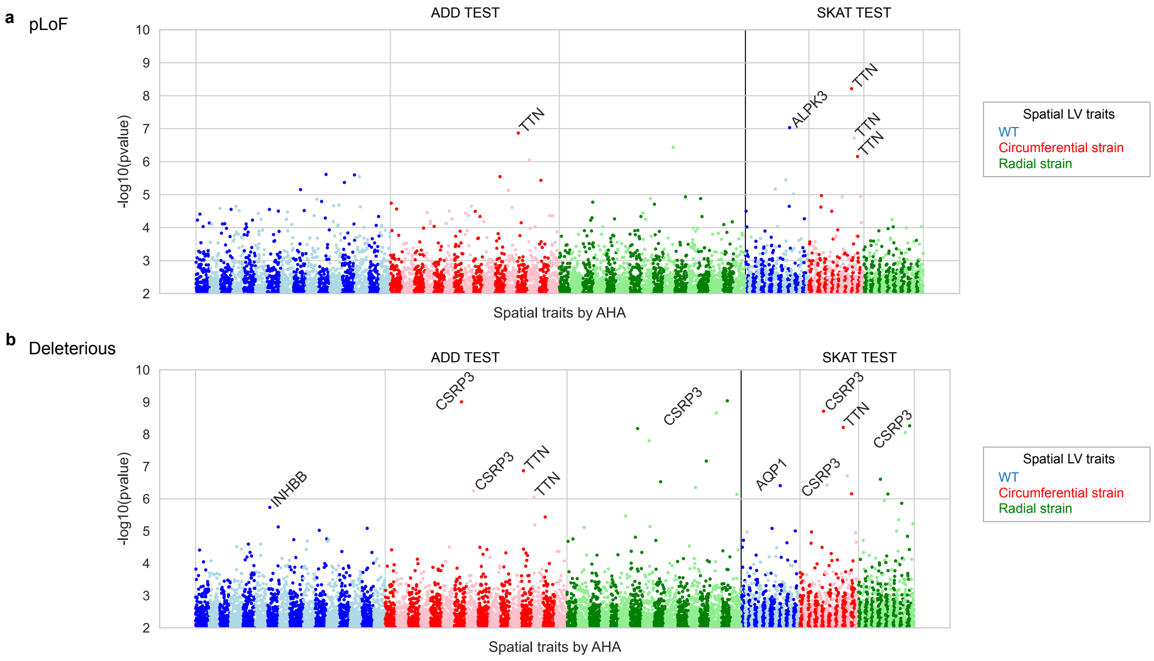


**Supplementary Figure 12. Exome-wide gene-based tests on spatial LV traits.** (a) Genes habouring more than five predicted loss-of-function (pLoF) variants in the cohort were tested for association with 48 spatial LV traits including wall thickness (blue), circumferential (red) and radial strain (green). Tests were performed using regular burden tests (ADD) and the variance component test (SKAT). ADD tests on pLoF variants were conducted for 10,768 genes, and SKAT for 3,666 genes. Alternating colors represent different AHA segments. Each dots represent the log-scaled P value of the gene association significance, and less genes were tested in SKAT. (b) Genes harbouring more than five predicted deleterious variants were tested. Similarly, 48 spatial LV traits were tested and alternating colors represent different AHA segments. ADD tests on pLoF variants were conducted for 15,924 genes, and SKAT for 9,732 genes. AHA: American Heart Association segment model. WT: wall thickness. SKAT: sequencing kernel association test.

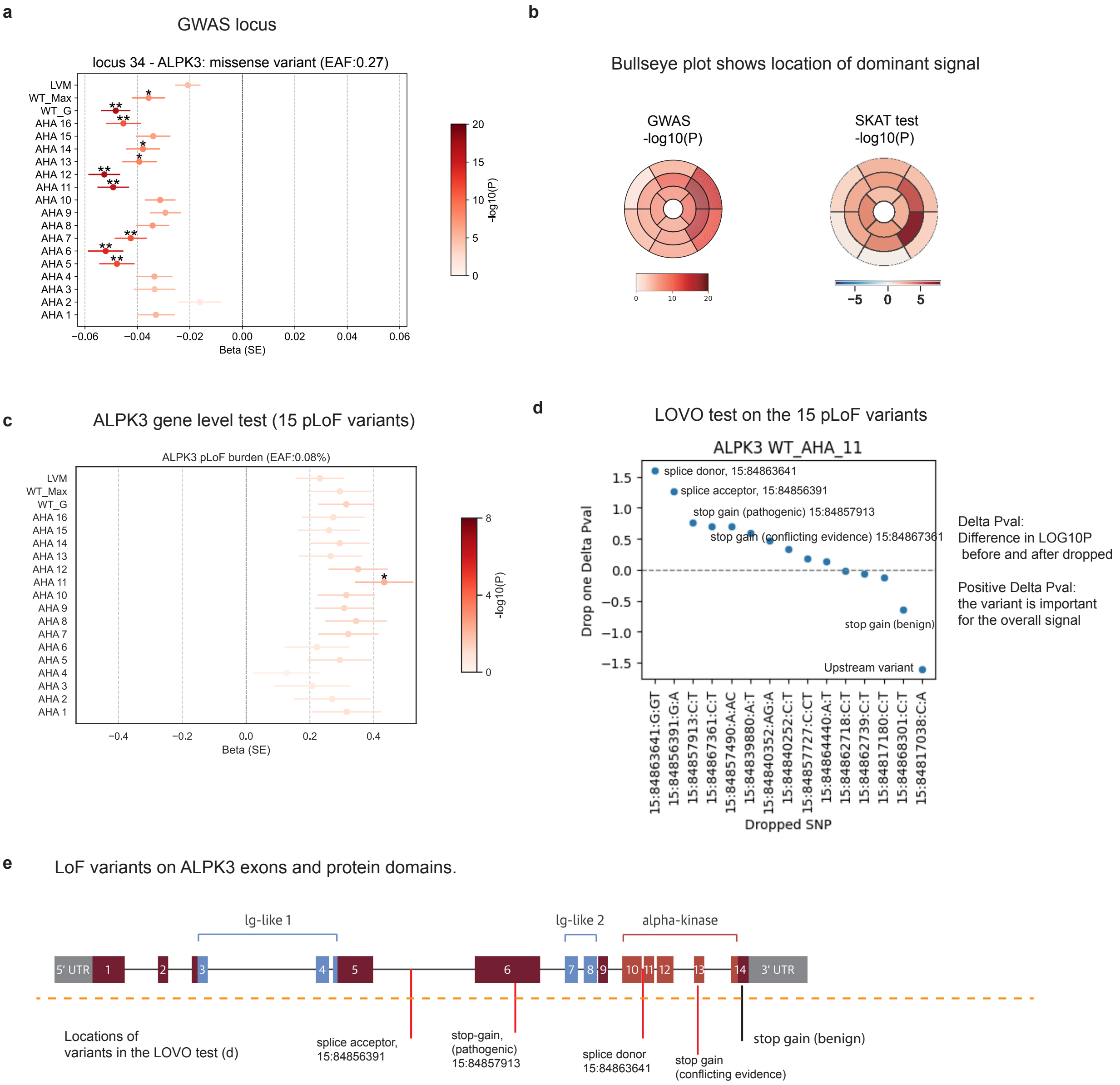


**Supplementary Figure 13. Combined GWAS lead and rare leave-one-variant-out analysis of ALPK3.** (a) Sentinel SNP in the GWAS locus and association values (beta) to global and spatial LV wall thickness, maximum wall thickness and LV mass. The minor allele is associated with decreased WT. (b) Bullseye plot that shows the P values of GWAS variant and SKAT burden tests. (c) The beta values of the ADD burdent tests, the pLoFs together were associated with increased WT. (d) Leave one variant out test on ALPK3, demonstraing different direction of effect from these variants. (e) the location of the variants on the ALPK3 domain structure. The gene scheme was duplicated from Almomani et.al., Figure 3[59]. The top four variants annotated in (d,e) from top to bottom are: splice donor rs753084997 (15:84863641), splice acceptor rs761330284 (15:84856391), stop gain (ClinVar pathogenic) rs749465164 (15:84857913), and stop gain rs541612157 (15:84867361).

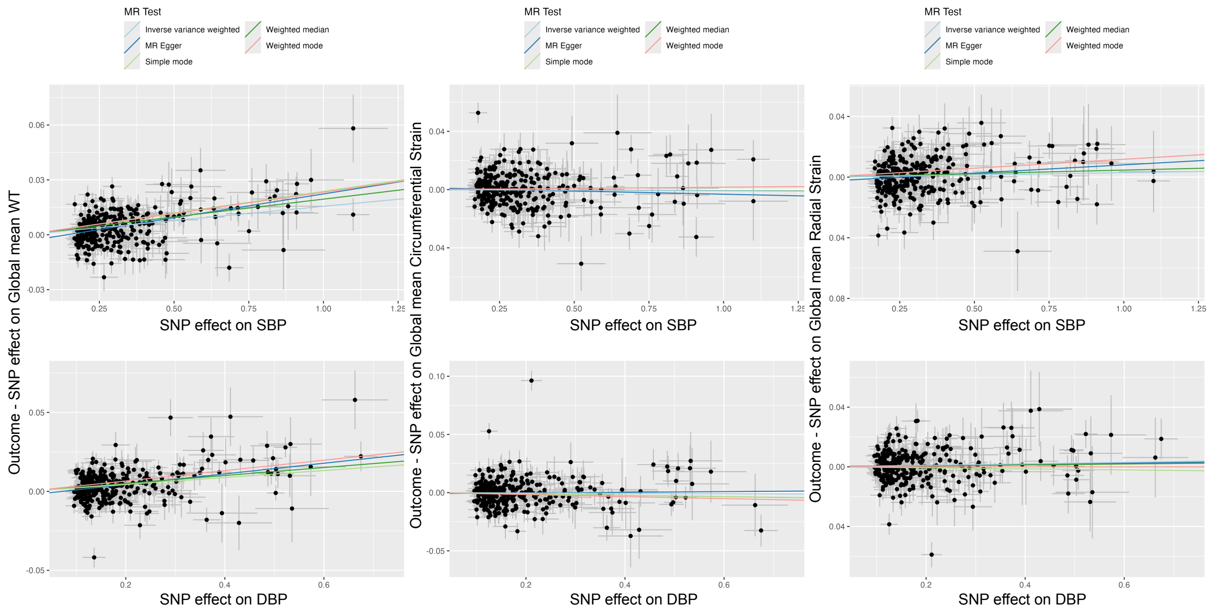


**Supplementary Figure 14. Single nucleotide polymorphism (SNP) effects of blood pressure on global LV traits**. Mendelian randomisation (MR) analysis of systolic (SBP) and diastolic (DBP) blood pressure as exposure (by row), global LV traits including the mean wall thickness, mean strain$^{circ}$ and mean strain$^{rad}$ as outcome (by column). Genetic instruments for SBP and DBP were selected from published GWAS[14]. The effects ($\beta$) of the exposure variable-increasing allele at independent SNPs ($r^2 < 0.001$) reaching $P < 5e\text{-}8$ are plotted as data points and associated standard errors are represented as lines extending from data points. The plots were produced using the R package `TwoSampleMR`. See Supplementary Data File 'Mendelian Randomisation.xlsx' for full MR results.

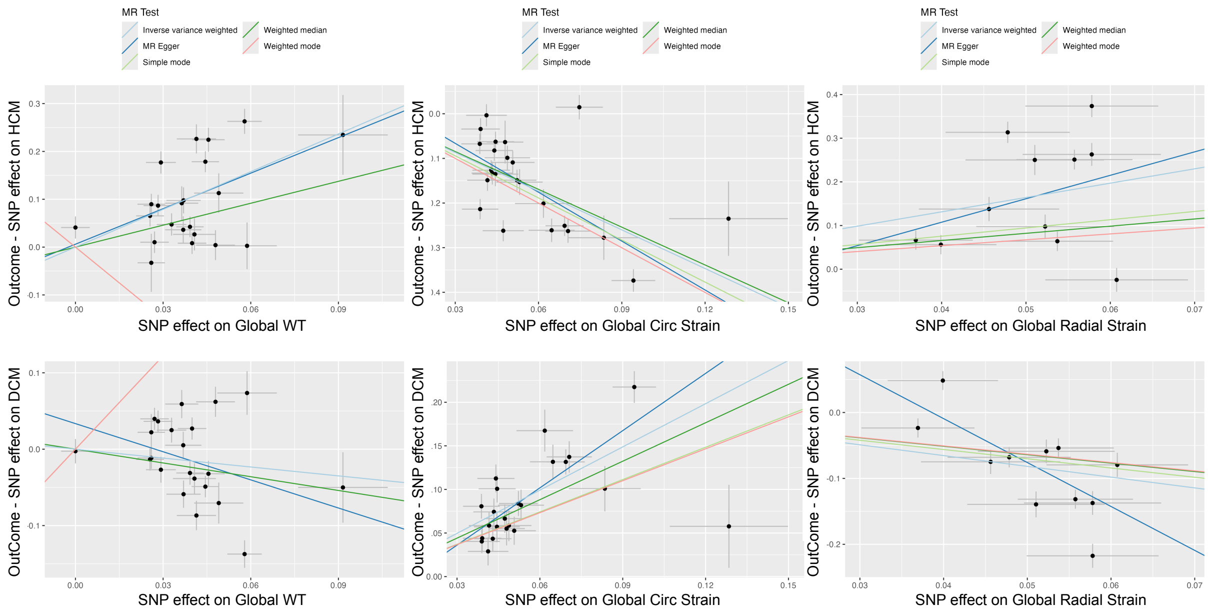


**Supplementary Figure 15. Single nucleotide polymorphism (SNP) effects of global LV traits on HCM and DCM**. Mendelian randomisation (MR) analysis of global LV mean wall thickness, mean strain$^{circ}$ and mean strain$^{rad}$ as exposure (by column), HCM and DCM as outcome (by row). The effects ($\beta$) of the exposure variable-increasing allele at independent SNPs ($r^2 < 0.001$) reaching $P < 5e\text{-}8$ are plotted as data points and associated standard errors are represented as lines extending from data points. Outcome were assessed with HCM[15] and DCM[16] GWAS. The plots were produced using the R package `TwoSampleMR`. See Supplementary Data File 'Mendelian Randomisation.xlsx' for full MR results.

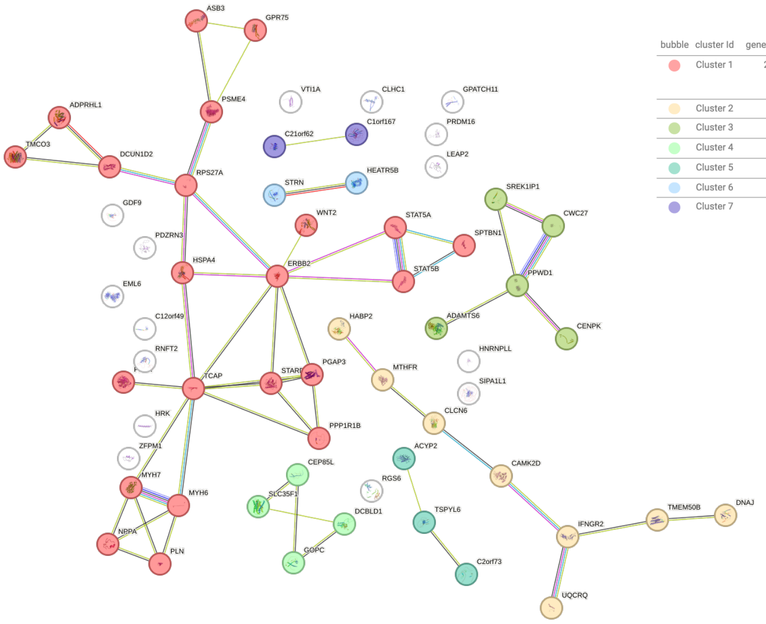


| bubble | cluster Id | gene count | protein names |
|---|---|---|---|
| | Cluster 1 | 22 | ADPRHL1, ASB3, DCUN1D2, ERBB2, GPR75, HSPA4, MYH6, MYH7, NPPA, PGAP3, PLN, PNMT, PPP1R1B, PSME4, RPS27A, SPTBN1, STARD3, STAT5A, STAT5B, TCAP, TMCO3, WNT2 |
| | Cluster 2 | 8 | CAMK2D, CLCN6, DNAJC28, HABP2, IFNGR2, MTHFR, TMEM50B, UQCRQ |
| | Cluster 3 | 5 | ADAMTS6, CENPK, CWC27, PPWD1, SREK1IP1 |
| | Cluster 4 | 4 | CEP85L, DCBLD1, GOPC, SLC35F1 |
| | Cluster 5 | 3 | ACYP2, C2orf73, TSPYL6 |
| | Cluster 6 | 2 | HEATR5B, STRN |
| | Cluster 7 | 2 | C1orf167, C21orf62 |

**Supplementary Figure 16. STRING analysis for protein coding genes in the spatial-only loci.** Protein coding genes from 18 spatial only GWAS loci were analysed using STRING database, k-means clustering identified seven clusters of interaction networks.

#### Supplementary Data Files

List of the Supplementary Data Files provided in the zip file.

- Supplementary Data File 1. LDSC Genetic Correlation full results.
- Supplementary Data File 2. GREML heritability and fixed effect variance full results.
- Supplementary Data File 3. PheWAS full results.
- Supplementary Data File 4. Spatial LV GWAS loci and gene prioritisation table.
- Supplementary Data File 5. Exome wide gene based burden tests significant associations.
- Supplementary Data File 6. Bidirectional Two-Sample Mendelian Randomisation full results.
- Supplementary Data File 7. Gene lists used in STRING-DB analysis and the list of enriched pathways.
